# Multi-ancestry meta-analysis of asthma identifies novel associations and highlights the value of increased power and diversity

**DOI:** 10.1101/2021.11.30.21267108

**Authors:** Kristin Tsuo, Wei Zhou, Ying Wang, Masahiro Kanai, Shinichi Namba, Rahul Gupta, Lerato Majara, Lethukuthula L. Nkambule, Takayuki Morisaki, Yukinori Okada, Benjamin M. Neale, Global Biobank Meta-analysis Initiative, Mark J. Daly, Alicia R. Martin

**Affiliations:** Analytic and Translational Genetics Unit, Massachusetts General Hospital, Boston, MA, USA; Stanley Center for Psychiatric Research, Broad Institute of MIT and Harvard, Cambridge, MA, USA; Program in Medical and Population Genetics, Broad Institute of MIT and Harvard, Cambridge, MA, USA; Department of Biomedical Informatics, Harvard Medical School, Boston, MA, USA; Department of Statistical Genetics, Osaka University Graduate School of Medicine, Suita, Japan; Howard Hughes Medical Institute and Department of Molecular Biology, Massachusetts General Hospital, Boston, MA USA; Department of Psychiatry and Mental Health, Faculty of Health Sciences, University of Cape Town, South Africa; Division of Molecular Pathology, The Institute of Medical Science, The University of Tokyo, Minato-ku, Japan; Laboratory for Systems Genetics, RIKEN Center for Integrative Medical Sciences, Yokohama, Japan; Laboratory of Statistical Immunology, Immunology Frontier Research Center (WPI-IFReC), Osaka University, Suita 565-0871, Japan; Department of Genome Informatics, Graduate School of Medicine, the University of Tokyo, Tokyo 113-0033, Japan; Center for Infectious Disease Education and Research (CiDER), Osaka University, Suita 565-0871, Japan; Institute for Molecular Medicine Finland, University of Helsinki, Helsinki, Finland

**Keywords:** asthma, GWAS, heterogeneity, multi-ancestry, polygenic risk prediction, cross-trait, subtypes

## Abstract

Asthma is a complex disease that affects millions of people and varies in prevalence by an order of magnitude across geographic regions and populations. However, the extent to which genetic variation contributes to these disparities is unclear, as studies probing the genetics of asthma have been primarily limited to populations of European (EUR) descent. As part of the Global Biobank Meta-analysis Initiative (GBMI), we conducted the largest genome-wide association study of asthma to date (153,763 cases and 1,647,022 controls) via meta-analysis across 18 biobanks spanning multiple countries and ancestries. Altogether, we discovered 179 genome-wide significant loci (p < 5×10^−8^) associated with asthma, 49 of which are not previously reported. We replicate well-known associations such as *IL1RL1* and *STAT6*, and find that overall the novel associations have smaller effects than previously-discovered loci, highlighting our substantial increase in statistical power. Despite the considerable range in prevalence of asthma among biobanks, from 3% to 24%, the genetic effects of associated loci are largely consistent across the biobanks and ancestries. To further investigate the polygenic architecture of asthma, we construct polygenic risk scores (PRS) using a multi-ancestry approach, which yields higher predictive power for asthma in non-EUR populations compared to PRS derived from previous asthma meta-analyses. Additionally, we find considerable genetic overlap between asthma age-of-onset subtypes, as well as between asthma and chronic obstructive pulmonary disease (COPD) but minimal overlap in enriched biological pathways. Our work underscores the multifactorial nature of asthma development and offers insight into the shared genetic architecture of asthma that may be differentially perturbed by environmental factors and contribute to variation in prevalence.

## Introduction

Asthma is a complex and multifactorial disease that affects millions of people worldwide, yet much of its genetic architecture has eluded discovery. Genetic factors contribute substantially to asthma risk, with heritability estimates from twin studies ranging between 50%-90%^1,2^. Early genome-wide association studies (GWAS) provided some evidence for the polygenic architecture of asthma^3–5^, but only in the past few years have genomic studies of asthma collated large enough sample sizes to more definitively articulate its polygenicity^6^. The most recent GWAS of asthma discovered 167 asthma-associated loci across the genome^7^. However, these risk loci only account for a small proportion of the total heritability of asthma. Furthermore, the discovery GWAS, like the majority of previous asthma GWAS, were primarily conducted in populations of European ancestry. Some major exceptions are the EVE Consortium^8^, one of the first efforts to perform GWAS in populations of African-American, African-Caribbean and Latino ancestries, as well as the Trans-National Asthma Genetic Consortium (TAGC)^9^ which included modest sample sizes from populations of African, Japanese and Latino ancestries in their meta-analysis. As these studies noted, efforts to conduct asthma GWAS in diverse populations are particularly important because the prevalence of asthma varies widely around the world. Surveys of asthma worldwide have found that prevalence can vary by as much as 21-fold among countries^10,11^. Within countries, prevalence of asthma ranges considerably as well^12^, and this variation cannot be attributed to any single known risk factor such as air pollution. Rather, the contributing genetic and environmental factors are complex. Therefore, assessing the genetic architecture of asthma in diverse cohorts is critical to gaining a more comprehensive understanding of asthma risk.

This heterogeneity in prevalence is mirrored by, and may be a consequence of, the heterogeneity of the disease itself. The wide variability in underlying mechanistic pathways and clinical presentations of asthma has led to a shift away from its characterization as a single disease entity^13–15^. Instead, asthma is now commonly viewed as a syndrome encompassing several distinct yet interrelated diseases, each driven by a unique set of genetic and non-genetic risk factors^13,14^. Different subgroups of asthma, for example, share genetic components with various comorbid diseases, including other respiratory diseases like chronic obstructive pulmonary disease (COPD), allergic diseases, obesity, and neuropsychiatric disorders^16–22^. This complexity in turn complicates standards for defining phenotypes to study; for example, one study found that nearly 60 different definitions of “childhood asthma” were used across more than 100 studies in the literature^23^. The heterogeneity of asthma thus presents many challenges in identifying genetic risk factors for asthma.

A greater understanding of the genetics underlying asthma risk can facilitate the development of more accurate clinical models of asthma that may help inform clinical intervention, prevention, and management strategies^24^. In particular, leveraging GWAS associations for genetic risk prediction models, such as polygenic risk scores (PRS), has shown potential in informing preventative clinical decision-making for several polygenic diseases^25–27^. For asthma, PRS could ultimately play a role in predicting disease severity and development in the clinical setting and serve as a tool for investigating gene-environment interactions in the research setting. So far, some GWAS have been applied to developing PRS for asthma^28–32^, but these models have had limited predictive ability, likely due to the insufficient sample sizes and diversity of existing datasets of asthma. This underscores the genetic complexity of asthma and highlights the need for more large-scale, genomic studies of asthma.

To more deeply interrogate the genetic architecture of asthma across different populations through genetic discovery and prediction, we analyzed paired phenotypic and genetic data from the Global Biobank Meta-analysis Initiative (GBMI). Participating biobanks shared summary statistics for the meta-analyses of 14 disease endpoints: asthma, COPD, heart failure, stroke, gout, venous thromboembolism, primary open-angle glaucoma, abdominal aortic aneurysm, idiopathic pulmonary fibrosis, thyroid cancer, cardiomyopathy, uterine cancer, acute appendicitis, and appendectomy^33^. More details on the selection of these disease endpoints can be found in Zhou et al. (2021)^33^. Compared to previous asthma resources and studies, this collaborative effort increased both the sample size and diversity of asthma cases by many folds, covered more variants with high imputation quality, and harmonized phenotypes using consistent electronic health record definitions for asthma across datasets. Harnessing this resource, we identify 49 loci not previously associated with asthma. Despite prevalence differences of nearly an order of magnitude, we also demonstrate remarkable consistency of genetic effects across the biobanks and ancestries in GBMI. Further, we show that the increased sample size and diversity of data from GBMI improves genetic risk prediction accuracy in multiple populations. Finally, we show that this meta-analysis captures much of the genetic architecture underlying asthma age-of-onset subtypes, and we provide additional evidence for shared genetic architectures between asthma and comorbid diseases such as COPD. Our findings highlight the need for future investigations into how genetic effects shared across different asthma subtypes and with different diseases contribute to the heterogeneity of asthma.

## Results

### Multi-ancestry meta-analysis for asthma across 18 biobanks in GBMI

To identify novel loci associated with asthma, we performed fixed-effects inverse-variance weighted meta-analysis using the harmonized GWAS summary statistics for asthma from 18 biobanks participating in GBMI (**Supplementary Table 1**). The combined sample size from all discovery studies was 153,763 cases and 1,647,022 controls, spanning individuals of European (EUR), African (AFR), Admixed American (AMR), East Asian (EAS), Middle Eastern (MID), and Central and South Asian (CSA) ancestry (**Fig. 1, Supplementary Fig. 1**). The meta-analysis of GWAS from four additional biobanks (9,991 cases and 63,605 controls) was used as an independent replication study (**Supplementary Table 1**). Despite the standardized phenotype definitions used by each biobank, which included the asthma PheCode and/or self-reported data (**Supplementary Table 3**), the prevalence of asthma varies widely across these biobanks, ranging from 3% in the Taiwan Biobank (TWB) to 24% in the Mass General Brigham Biobank (MGB). We applied pre- and post-GWAS quality control filters that resulted in 70.8 million single-nucleotide polymorphisms (SNPs) for meta-analysis; for downstream analyses we analyzed SNPs present in at least 2 biobanks^33^. The meta-analysis identified 179 loci of genome-wide significance (p < 5×10^−8^), 49 of which have not been previously reported to be associated with asthma (**Fig. 2A, Supplementary Fig. 2**). These potentially novel loci were defined so that the index variants, or the most significant variants in each locus, were at least 1 Megabase in distance from a previously discovered genome-wide significant variant associated with asthma (**Methods**). Additionally, all but one index variant did not have a previously discovered SNP in linkage disequilibrium (LD) at *r*^*2*^ > 0.07, estimated using a reference panel from individuals in 1000 Genomes^34^ (**Supplementary Table 2**). In the replication meta-analysis, 51 of the 179 loci had index variants with a p-value < 0.05, even though the case numbers in the replication data were less than 10% of the case numbers in the discovery data (**Supplementary Table 2**). 154 of the 179 index variants had consistent directions of effect in the discovery and replication meta-analyses. We also found that the potentially novel associations had smaller effect sizes on average compared to the previously reported loci, across the allele frequency spectrum (**Fig. 2B**). This illustrates that with the increased power and effective sample size of GBMI, we were able to uncover SNPs with more modest effects on asthma.

**Figure 1.**
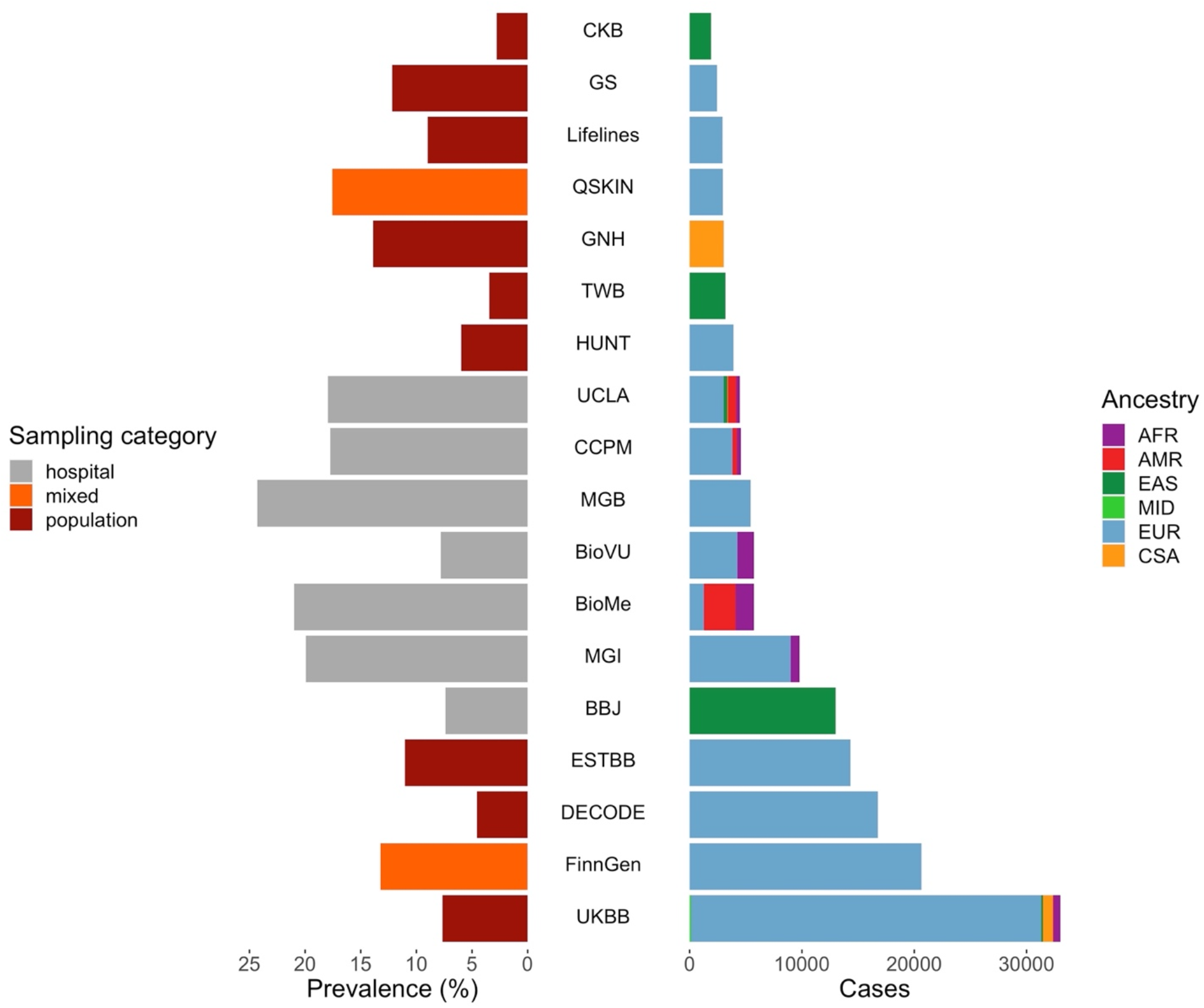
18 biobanks in GBMI contributing GWAS of asthma. Distribution of prevalence of asthma on left and number of cases of asthma on right across biobanks in GBMI. Biobanks span different sampling approaches and ancestries (AFR = African; AMR = Admixed American; EAS = East Asian; MID = Middle Eastern; EUR = European; CSA = Central and South Asian).

**Figure 2.**
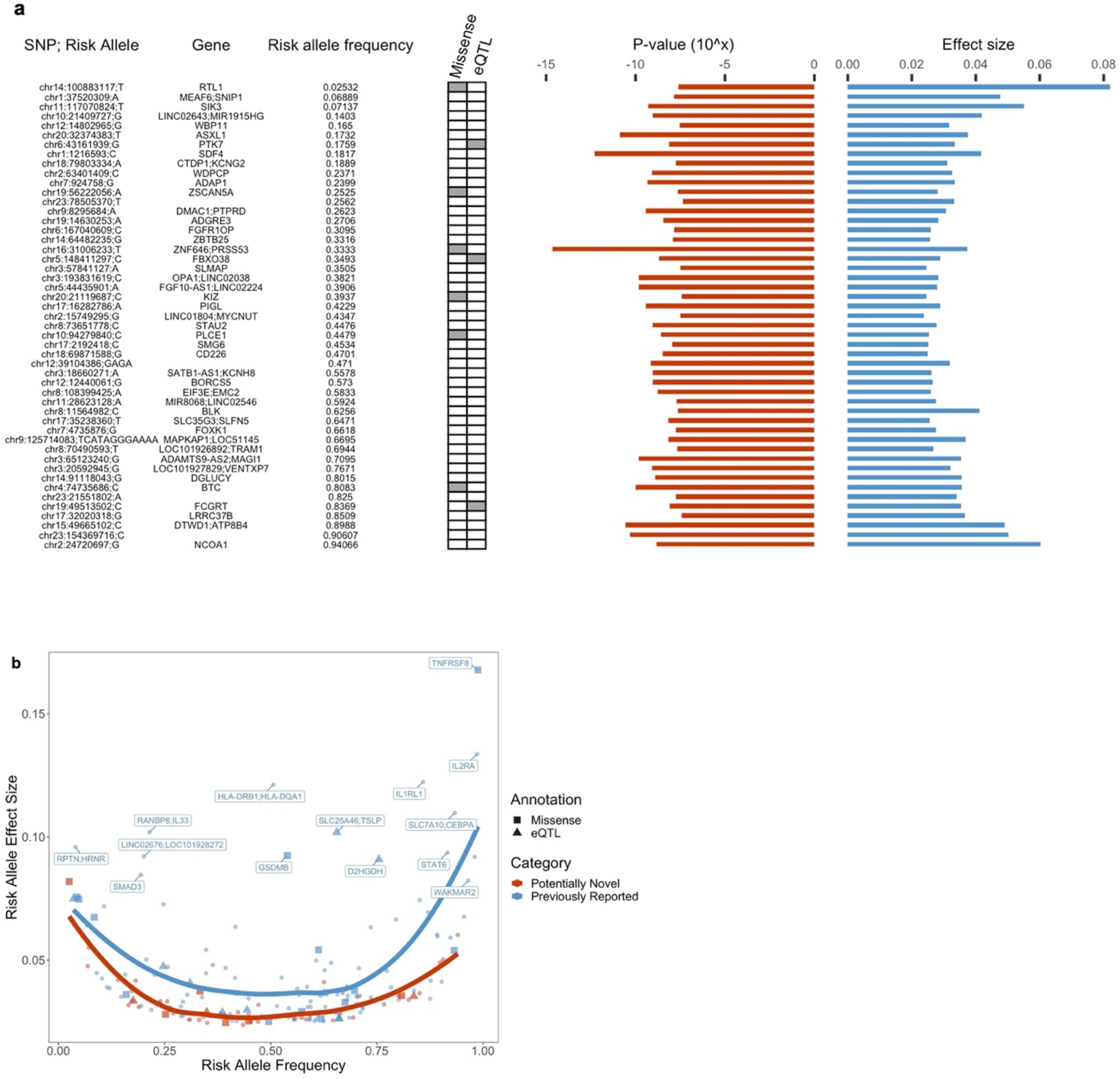
Top loci associated with asthma. **a**, Index variants of 49 asthma-associated loci that are potentially novel. Missense variants and cis-eQTLs fine-mapped with PIP > 0.9 that overlapped with an index or tagging variant (*r*^*2*^ > 0.8) are annotated here. Frequency of risk allele and effect size estimate in GBMI meta-analysis are shown on the right. **b**, Frequency and effect size of risk alleles of all 179 index variants. Previously reported genes with large effect sizes are highlighted.

Because the GBMI meta-analysis includes data from UKBB, we compared our results to the TAGC meta-analysis results that did not include the UKBB GWAS to facilitate analyses that require independent samples^9^. Of the index variants within the top 179 loci in GBMI, 122 were in the TAGC meta-analysis or had a tagging variant in high LD (*r*^*2*^ > 0.8) in the TAGC study; 76 of these had p < 0.05 in the TAGC results. We compared the effect sizes of these 76 SNPs in the GBMI and the TAGC meta-analyses using a previously proposed Deming regression method that considers standard errors in both effect size estimates^35^. We found that all 76 SNPs were directionally consistent and aligned across the studies (**Supplementary Table 4, Supplementary Fig. 3**).

Among the 49 novel loci, the index variants of six loci were missense or in high LD (*r*^*2*^ > 0.8) with a missense variant (**Supplementary Table 2**). One of these SNPs, chr10:94279840:G:C (p_meta-analysis_ = 2.5×10^−9^), resides in *PLCE1*, an autosomal recessive nephrotic syndrome gene^36^; high prevalence of atopic disorders, like asthma, among children with nephrotic syndrome has long been observed in the clinic, suggesting potential shared pathways underlying asthma and nephrotic syndrome^37^. The asthma risk allele has also been previously linked to lower blood pressure^38^. The index SNPs chr14:100883117:G:T (p_meta-analysis_ = 2.6×10^−8^) and chr19:56222056:C:A (p_meta-analysis_ = 2.4×10^−8^) also implicate novel genes, *RTL1* and *ZSCAN5A* respectively. *RTL1* has been found to play a role in muscle regeneration^39^, and *ZSCAN5A* has been linked to monocyte count^40^. Additionally, three of the novel index SNPs co-localized with a fine-mapped cis-eQTL (**Supplementary Table 2**). One of these, chr19:49513502:C:T (p_meta-analysis_ = 7.98×10^−9^), implicates *FCGRT*, which regulates IgG recycling and is a potential drug target for autoimmune neurological disease therapies^41^. The other previously-reported missense variants replicated previous findings; among these, chr4:102267552:C:T (p.Ala391Thr, p = 2.5×10^−12^) is a highly pleiotropic variant in *SLC39A8* that has been associated with many psychiatric, neurologic, inflammatory and metabolic diseases^42–48^ and has been demonstrated to disrupt manganese homeostasis^49^. Variants implicating well-known asthma-associated genes with large effects, like *IL1RL1, IL2RA, STAT6, IL33, GSDMB*, and *TSLP*, were replicated in the meta-analysis as well.

### GWAS from diverse ancestries reveal shared genetic architecture of asthma and improves power for genetic discovery

Given that sample size, disease prevalence, ancestry, and sampling approaches differed across the 18 biobanks, we investigated the consistency of the asthma-associated loci across the biobanks and their attributes. We first implemented an approach that estimates the correlation (*r* _*b*_) between the effects of the index variants of the 179 top loci in each biobank GWAS and the corresponding meta-analysis excluding that biobank^50^. We observed that most biobanks have highly correlated genetic effects with other biobanks (*r* _*b*_ estimates close to 1) (**Supplementary Table 5**). To further interrogate the consistency of the index variants in all biobanks, we computed the ratio of the effect size of these SNPs in the biobank-specific GWAS over that in the corresponding leave-that-biobank-out meta-analysis. We found that the average per-biobank ratios were almost evenly split between those greater than and less than 1 (**Supplementary Fig. 4**). This indicates that any significant difference in effects likely does not reflect technical artifacts in the meta-analysis or GWAS procedures. We also applied Deming regression^35^ to assess the alignment of the SNP effects in each biobank-specific GWAS with the effects in the corresponding leave-that-biobank-out meta-analysis and observed that the effect sizes were comparable across the biobanks (**Fig. 3, Supplementary Fig. 5**). Furthermore, the genome-wide genetic correlations between the biobanks with non-zero heritability estimates and the respective leave-that-biobank-out meta-analyses were all close to 1^33^.

**Figure 3.**
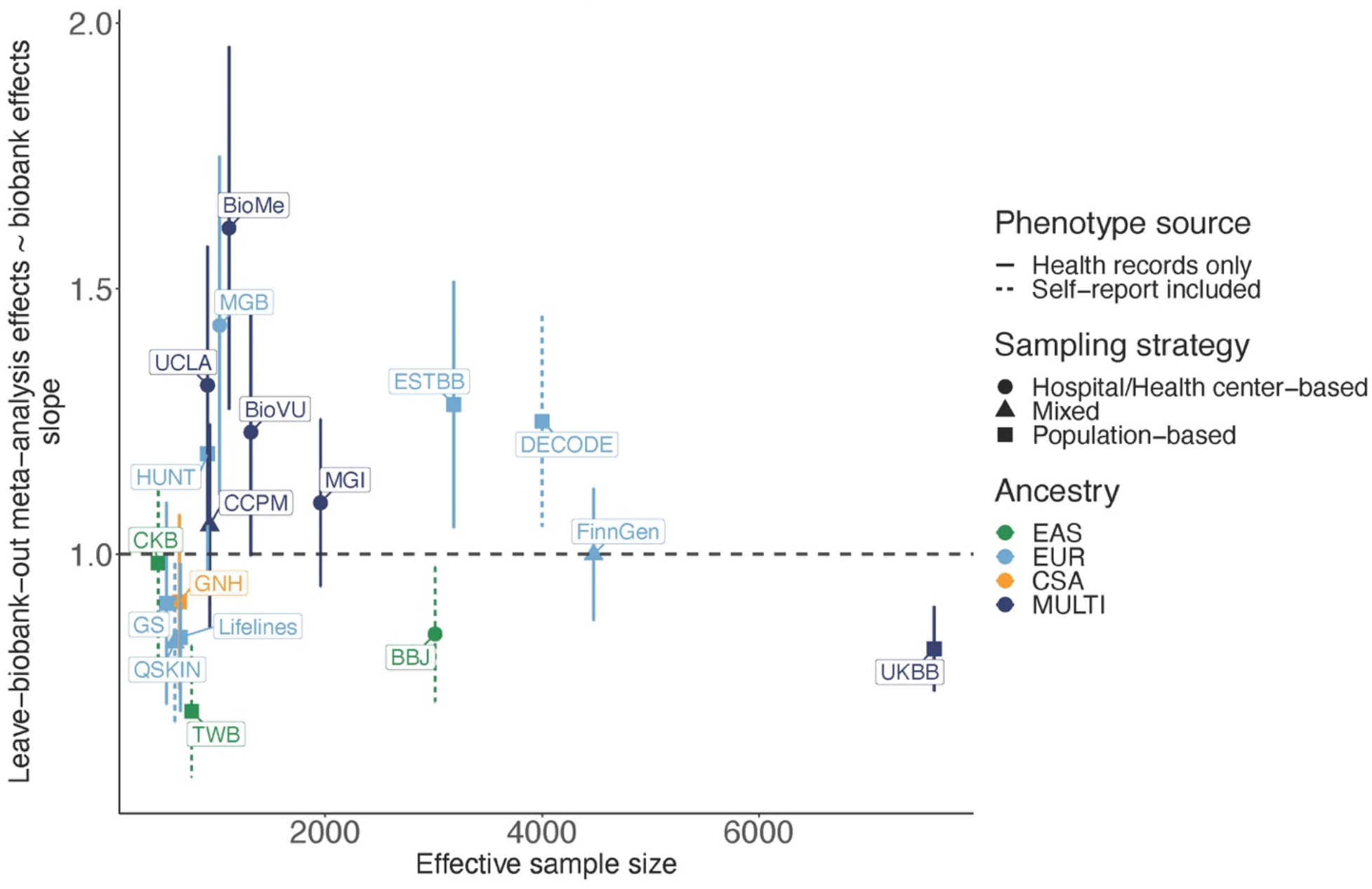
Consistency of loci across biobanks. Regression slopes computed using the Deming regression method, which compared effects of index variants in each biobank GWAS against their effects in the corresponding leave-that-biobank-out meta-analysis^23^. The x-axis shows the effective sample size of each biobank, computed as 4/(1/cases + 1/controls).

To test for potential heterogeneity in effect estimates due to ascertainment, we conducted an additional sensitivity analysis comparing SNP effects in the meta-analyses of the hospital-vs. population-based biobanks. We conducted meta-analyses of the 9 population-based biobanks (CKB, DECODE, ESTBB, GNH, GS, HUNT, Lifelines, TWB, and UKBB) and 6 hospital-based biobanks (BBJ, BioMe, BioVU, MGB, MGI, and UCLA). We then fitted the Deming regression^35^ on the effect size estimates of the loci identified by the all-biobank meta-analysis, using the SNPs with p-value < 1×10^−6^ in both meta-analyses, and observed high consistency in the effects across the two groups (**Supplementary Fig. 6**).

Taken together, these analyses indicate that the genetic architecture of asthma is largely shared across cohorts, despite differences in characteristics like disease prevalence and ascertainment strategy. Furthermore, the consistency of genetic effects across the biobanks suggests that the fixed effects meta-analysis approach is appropriate for the integration of GWAS from the different datasets. We additionally conducted meta-analysis using the meta-regression approach implemented in MR-MEGA^51^, which accounts for potential effect size heterogeneity across datasets. MR-MEGA identified only 2 additional loci associated with asthma, 1 of which is novel (**Supplementary Table 6**).

We also found little evidence of heterogeneity in the ancestry-specific effect sizes for the index variants. One SNP, chr10:9010779:G:A, was significantly heterogeneous (p-value for Cochran’s Q test < 0.0003, the Bonferroni-corrected p-value threshold) across the ancestry-specific meta-analyses of AFR, AMR, CSA, EAS, and EUR individuals (**Fig. 4A, Supplementary Table 7**). One known SNP that nearly reached the Bonferroni-corrected p-value threshold for heterogeneity, chr16:27344041:G:A, displayed different effects in the EUR and EAS cohorts. This SNP lies within an intron of *IL4R* (**Fig. 4B**), which has known associations with asthma^6,52^. Previous studies have investigated the association of *IL4R* with asthma in different populations, with inconsistent results, so future studies on the population-specific effects of this gene will be needed^53–55^. Our findings demonstrate that despite broad consistency of effect sizes across ancestries among the top loci, the increased power and diversity of GBMI enabled the detection of SNPs with significantly different effects across ancestries.

**Figure 4.**
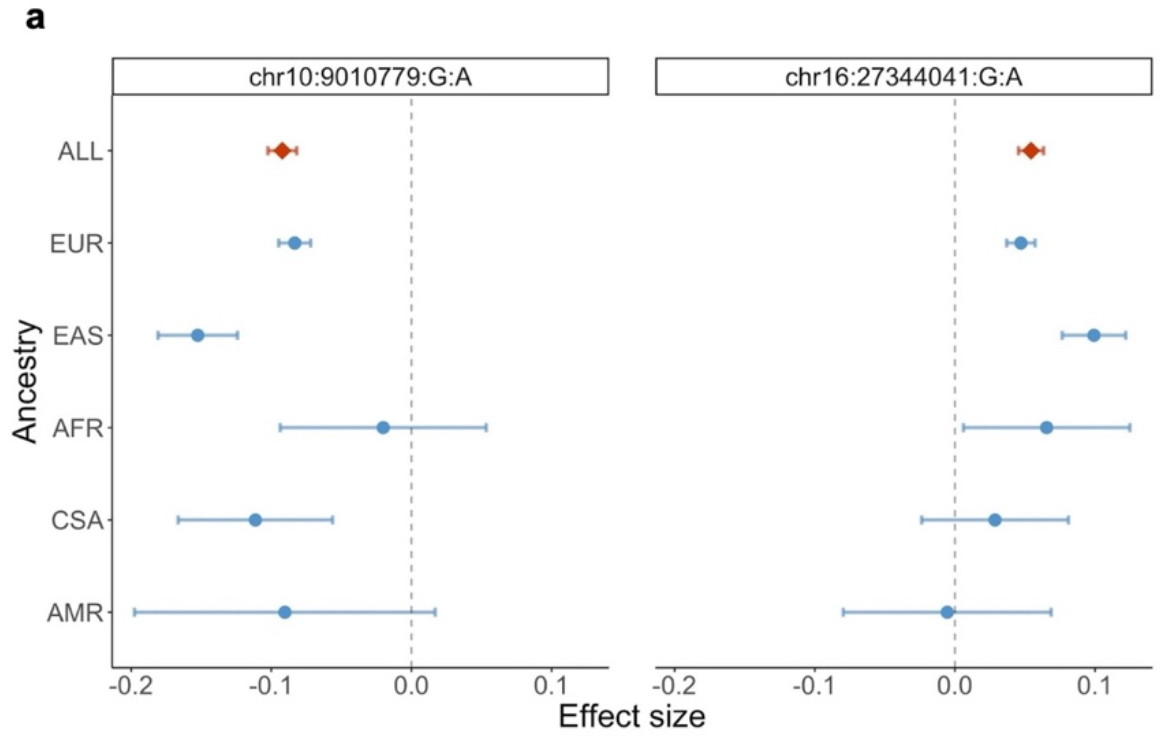
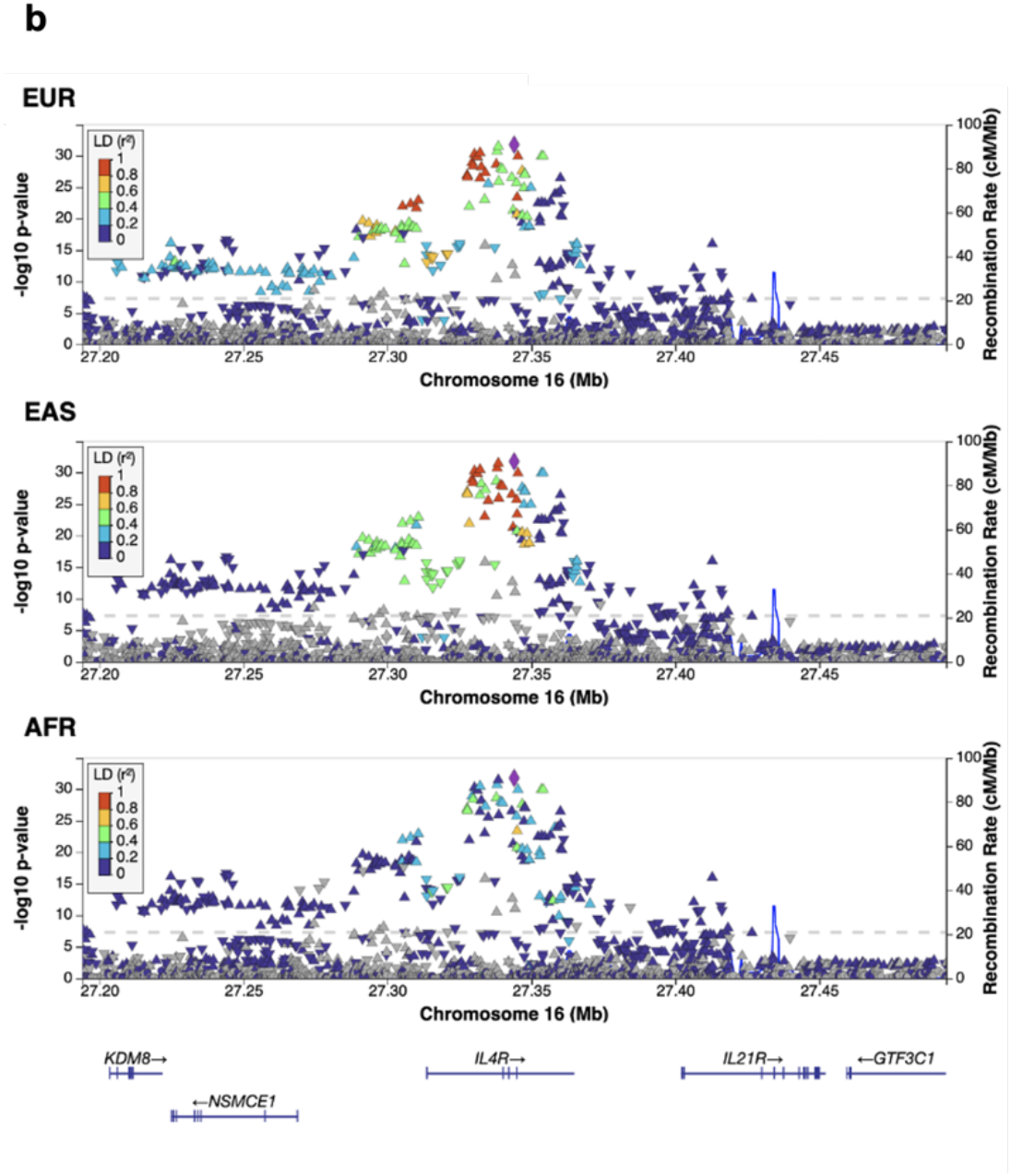
Loci showing heterogeneity in ancestry-specific effect sizes. **a**, The index variants with the most significant p_Cochran’s Q_. Effect sizes of these variants in each ancestry-specific meta-analysis are shown here. **b**, LocusZoom plots showing the association of chr16:27344041:G:A (purple symbol) and variants within 150kb upstream and downstream of this variant with asthma. Color coding of other SNPs indicates LD with this SNP. EUR, EAS, and AFR indicate the population from which LD information was estimated.

Additionally, the greater diversity of GBMI facilitated the discovery of loci that would not have been identified in association analyses using data from only EUR ancestry cohorts. We found that of the 179 loci identified in the all-biobank meta-analysis, 49 did not reach genome-wide significance in the EUR-only meta-analysis (**Supplementary Table 8**). This additional yield of loci may be partially due to the increase in sample size, but the inclusion of GWAS from diverse ancestries also enabled the identification of loci that are more frequent in some non-EUR populations. 19 of these 49 loci were potentially novel, and 13 of these novel loci had an index variant higher in frequency in a non-EUR ancestry group compared to the EUR ancestry group. The consistent effect estimates of the 49 additional variants across populations (45/49 had p-value for Cochran’s Q test across ancestries >0.02) indicate that the additional variants discovered with the incorporation of GWAS from diverse ancestries do not tend to be population-specific loci that only have effects in certain populations. However, due to differences in frequency across populations, it is essential to conduct asthma GWAS in different populations to uncover the full spectrum of asthma-associated loci.

### Meta-analysis across diverse ancestries improves asthma PRS accuracy

We next explored the impact of the increased sample sizes and diversity in GBMI on genome-wide risk prediction of asthma. To establish a baseline understanding of PRS performance for asthma as well as other disease endpoints in GBMI, Wang et al. (2021)^56^ evaluated and compared the prediction accuracy of PRS derived from the pruning and thresholding (P+T) method and PRS-CS^57^ in target cohorts of EUR, CSA, EAS, and AFR ancestries, using the leave-one-biobank-out meta-analyses as discovery data. This study observed improvements in prediction accuracy for asthma using PRS-CS across all target cohorts (**Supplementary Fig. 7**), and additionally, the PRS derived from the GBMI leave-one-biobank-out meta-analyses of asthma had higher predictive accuracy, as measured by *R*^*2*^ on the liability scale 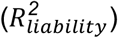, compared to the PRS constructed from the TAGC meta-analysis^9^ (**Fig. 5**).

**Figure 5.**
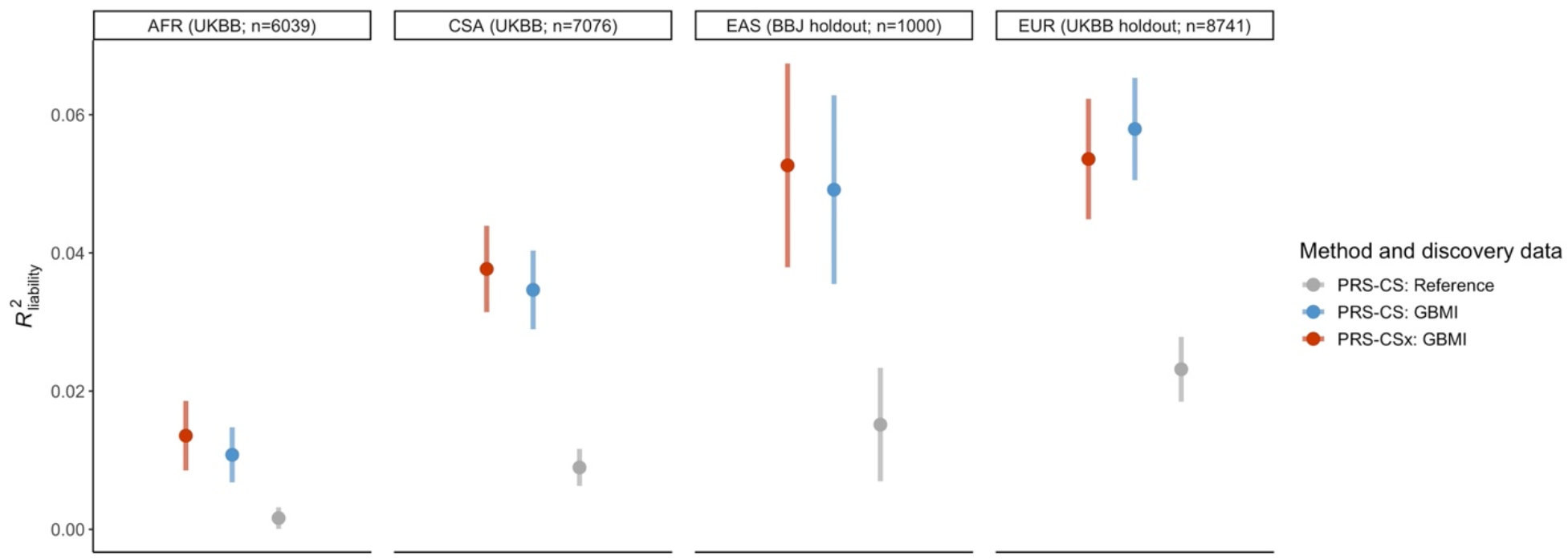
PRS performance across ancestries. Each panel represents a target cohort in which PRS constructed using PRS-CSx and PRS-CS were evaluated. PRS-CS analyses used the GBMI leave-BBJ-out meta-analysis and GBMI leave-UKBB-out meta-analysis as discovery data for the BBJ and all UKBB target cohorts, respectively (Supplementary Table 10)^45^. The reference dataset was the TAGC meta-analysis^5^. Sample sizes for the target cohorts are: cases=849 and controls=5190 for AFR; cases=500 and controls=500 for EAS; cases=1164 and controls=7577 for EUR; cases=1232 and controls=6744 for CSA. Error bars represent standard deviation of R^2^ on the liability scale across 100 replicates.

To expand on these analyses, we tested a recently-developed extension of PRS-CS, PRS-CSx^58^, for asthma risk prediction. This method jointly models multiple summary statistics from different ancestries to enable more accurate effect size estimation for prediction. For input to PRS-CSx, we used the AFR, AMR, EAS, CSA, and EUR ancestry-specific meta-analyses from GBMI; the discovery meta-analysis that matched the ancestry of the target cohort excluded the target cohort (**Supplementary Fig. 8**). With the posterior SNP effect size estimates from PRS-CSx, we tested the multi-ancestry PRS in the following target populations: AFR ancestry individuals in UKBB, CSA ancestry individuals in UKBB, a holdout set of EAS ancestry individuals in BBJ, and a holdout set of EUR ancestry individuals in UKBB. The final prediction models tested in these target populations were the optimal linear combinations of the population-specific PRS. The average 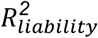 in the EAS (0.053) and EUR (0.054) target cohorts approached the SNP-based heritability 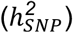, estimated to be 0.085 for asthma using the all-biobank meta-analysis^56^, while the prediction accuracies in the CSA (0.038) and AFR (0.014) target cohorts were lower (**Fig. 5, Supplementary Table 9**). When we downsampled the EUR target cohort to 1,000 individuals, to match the sample size of the EAS target cohort, we found a higher average 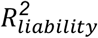 (0.063) but, as expected, much larger confidence intervals (**Supplementary Fig. 9**). 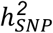 estimates may differ across biobanks and ancestries given differences in disease prevalence, environmental exposures, phenotype definitions, and other factors, and these differences may in turn contribute to the PRS in EAS individuals performing similarly to PRS in EUR individuals in our analyses, despite the smaller sample size of the EAS discovery cohort. The 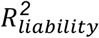 across the target populations for the PRS-CSx scores were roughly the same as the 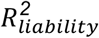 of the PRS derived from the PRS-CS analyses. It is important to note that the discovery data used in the PRS-CS analyses differed slightly in sample size and composition, since the leave-one-biobank-out approach was used for PRS-CS, but the target cohorts in which the PRS were evaluated were the same (**Supplementary Table 10**).

To investigate why improvement in performance using PRS-CSx was only incremental in most of the target cohorts, we examined the performances of each population-specific PRS. We found that across all target cohorts, PRS derived from either the EUR or EAS set of posterior effect size estimates outperformed the linear combination, and the 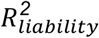 of these PRS were also higher compared to that of the PRS-CS scores (**Supplementary Fig. 10, Supplementary Table 9**). This suggests that the addition of more discovery GWAS to PRS-CSx can improve the accuracy of PRS based on a single set of posterior effect size estimates, but the linear combination of PRS from multiple GWAS does not necessarily yield higher accuracy. This may be due to the considerably smaller sample sizes of some of the input discovery meta-analyses in our analyses and thus varying signal to noise ratios. Collectively, these analyses show that the increase in scale and diversity of discovery GWAS for PRS is the primary driver of increased PRS accuracy in non-EUR populations for asthma, with marginal gains using PRS-CSx over PRS-CS. For EUR target cohorts, a multi-ancestry PRS construction method like PRS-CSx does not seem to contribute much improvement in prediction accuracy, likely due to the predominating sample size of EUR discovery GWAS, as well as the inclusion of GWAS from smaller, non-EUR discovery cohorts which may introduce more noise than signal.

### Childhood-onset (COA) and adult-onset (AOA) asthma are highly genetically correlated

To increase power for genetic discovery, we used a broad phenotype definition for asthma (**Methods**), but given the heterogeneity of the disease, we sought to address the extent to which this meta-analysis captured the genetic architectures of two common subtypes of asthma, childhood-onset (COA) and adult-onset (AOA) asthma. We conducted asthma age-of-onset subtype analyses in two of the participating GBMI biobanks for which age at asthma diagnosis information were accessible, UKBB and FinnGen. Using a cut-off age of 19 years at asthma diagnosis to define the subtypes (**Methods**), we performed GWAS of COA and AOA in FinnGen and the EUR ancestry cohort in UKBB, as well as fixed-effects, inverse-variance weighted meta-analyses of the COA (20,964 cases, 674,014 controls) and AOA (56,744 cases, 674,014 controls) GWAS, respectively. Applying linkage-disequilibrium score correlation (LDSC), we observed strong genetic correlations between each COA GWAS and the respective leave-that-biobank-out meta-analysis of all other biobanks utilizing the broad phenotype definition (*r*_*g*_ (se) *=* 0.73 (0.03), p = 4.70×10^−132^ for UKBB and *r*_*g*_ (se) *=* 0.80 (0.4), p = 3.19×10^−73^ for FinnGen), and even larger genetic correlations between each AOA GWAS and leave-that-biobank-out meta-analysis (*r*_*g*_ (se) *=* 0.90 (0.04), p = 1.71×10^−127^ for UKBB and *r*_*g*_ (se) *=* 0.90 (0.30), p = 1.39×10^−237^ for FinnGen). The genetic correlation between the COA and AOA meta-analyses was similarly high (*r*_*g*_ (se) *=* 0.78 (0.30), p = 1.32×10^−116^), and similar to the genetic correlation (*r*_*g*_ (se) *=* 0.67 (0.02)) reported by a previous study of asthma age-of-onset subtypes^59^. We also observed substantial overlap between the top loci identified in each subtype meta-analysis and the all-asthma meta-analysis. 75 of the 90 loci (83%) of genome-wide significance (p < 5×10^−8^) and 55 of the 69 loci (80%) identified by the COA and AOA meta-analysis, respectively, overlapped with a locus discovered in the all-asthma meta-analysis (**Supplementary Table 11**). Overall, these results suggest that much of the genetic architecture between COA and AOA is shared, as is consistent with previous findings^59,60^. Despite the GBMI meta-analysis drawing from primarily adult cohorts, many of the genetic variants identified contribute to both subtypes.

To investigate whether the genetic effects of the index variants of the asthma-associated loci differ across the subtypes, we compared the estimated effect sizes of the 179 index variants discovered in the all-asthma meta-analysis in the COA and AOA meta-analyses using the Deming regression method. We found that these variants had systematically stronger effects in the COA meta-analysis compared to in the AOA meta-analysis (**Supplementary Fig. 11**), supporting previous findings that the etiology of COA is likely partially characterized by genes that have smaller (or no) effects on AOA^59,60^.

### Asthma and COPD have shared and distinct biological processes

The shared genetic factors between asthma and different diseases that often coexist with asthma, such as COPD, a late-onset respiratory disease, have also been used to investigate and characterize asthma heterogeneity. It has been well-documented in the literature that asthma and COPD are frequent comorbidities of each other^61^, but only a few studies thus far have investigated the extent to which this is driven by a shared genetic basis^62–64^. Utilizing the GBMI meta-analyses of asthma and COPD, we observed a strong genetic correlation between asthma and COPD (*r*_*g*_ (se) = 0.67 (0.021), p = 1.55×10^−226^). This genetic correlation estimate is higher than estimates from previous studies, which ranged from 0.38-0.42^63,64^. This may be a result of the discovery datasets used by these studies, which were enriched for pediatric asthma cohorts, while GBMI biobanks are primarily composed of adult participants. To more formally test for potential differences in the shared genetic architecture of age-of-onset subtypes and COPD, we computed genetic correlations between the COA and AOA meta-analyses and the GBMI COPD meta-analysis. We found that the AOA meta-analysis had a strong genetic correlation with the COPD meta-analysis (*r*_*g*_ (se) *=* 0.60 (0.3), p = 2.65×10^−94^), while the COA meta-analysis had a more moderate genetic correlation with the COPD meta-analysis (*r*_*g*_ (se) *=* 0.33 (0.3), p = 7.60×10^−31^).

To further evaluate the extent of genetic overlap between asthma and COPD, we applied a gene prioritization method, Multi-marker Analysis of GenoMic Annotation (MAGMA)^65^, to the GBMI EUR, AFR, EAS, and CSA meta-analyses of asthma as well as the GBMI EUR, AFR, and EAS meta-analyses of COPD. After Bonferroni correction, we found that 442, 149, and 6 genes were significantly associated with asthma in the EUR (p < 2.50×10^−6^), EAS (p < 2.50×10^−6^), and CSA (p < 2.52×10^−6^) populations, respectively, with no significantly associated genes in the AFR cohort (all p > 2.51×10^−6^) (**Supplementary Table 14**). The majority of the genes associated with asthma identified in the EAS meta-analysis overlapped with the genes from the EUR meta-analysis (126 out of 149 genes), and all 6 genes associated with asthma as identified in the CSA meta-analysis were also significantly associated in the EUR and EAS meta-analyses. We identified 46 and 33 genes significantly associated with COPD in the EUR (p < 2.50×10^−6^) and EAS (p < 2.50×10^−6^) cohorts, respectively, and similarly to asthma, no significantly associated genes from the AFR meta-analysis (all p > 2.51×10^−6^) (**Supplementary Table 15**). Of the 75 genes associated with COPD across the EUR and EAS meta-analyses, 24 overlapped with the asthma-associated genes. We also conducted gene prioritization using Data-driven Expression-Prioritized Integration for Complex Traits (DEPICT)^66^ and gene-level Polygenic Priority Score (PoPS)^67^. However, only 3 of the 52 genes (6%) prioritized for COPD by DEPICT overlapped with a gene prioritized for asthma using the same method (**Supplementary Table 16**), and 17 of the 184 genes (9%) prioritized for COPD by PoPS overlapped with a prioritized gene for asthma (**Supplementary Table 17**). Across the shared COPD and asthma genes prioritized by each method, only 1 gene, *MED24*, was prioritized by more than one method, highlighting that existing gene prioritization methods have poor agreement, an observation that has been previously discussed^67^ and is explored in more detail in Zhou et al. (2021)^33^.

We also adopted MAGMA for gene-set enrichment based on the curated and ontology gene sets from the Molecular Signatures Database (MSigDB)^68^. We found hundreds of gene sets that were significantly enriched (FDR < 0.05) by the asthma-associated genes discovered in the EUR and EAS meta-analyses (**Supplementary Table 18**). In contrast, only a handful of gene sets were significantly enriched among COPD-associated genes discovered in the AFR meta-analysis, likely reflecting the smaller overall sample size of the COPD meta-analysis (**Supplementary Table 19**). The top-ranked asthma pathways from the EUR meta-analysis included cytokine and interleukin signaling and T-cell activation. Consistently biologically, the EAS meta-analysis identified autoimmune thyroid disease and graft vs. host disease pathways. The top-ranked COPD pathways from the EUR meta-analysis, although not significant, included several pathways related to nicotine receptor activity. These results reinforce that despite the substantial genetic overlap, asthma and COPD are governed by distinct biological processes as well. Future investigations will be required to fully parse out the etiology and comorbidities of asthma, like COPD, that develop later on in adulthood.

### Genetic overlap between asthma and other diseases

Non-genetic epidemiological studies have also identified correlations between asthma and many other disease categories beyond COPD^69–71^. More recently, some genome-wide cross-trait studies have found evidence for shared genetic architectures between asthma and other allergic diseases^21,72^, neuropsychiatric disorders^22^, and obesity^20^, suggesting that a comprehensive characterization of the shared genetics among asthma and other complex diseases and traits could provide insights into the variable pathology of asthma^19^. Together, these findings motivated us to assess whether correlations across a broad spectrum of disease endpoints are potentially driven by a shared genetic basis, or are purely observational and not driven by a shared biology. Since the GBMI project was limited to 14 disease endpoints, we utilized the wide range of phenotypic data available in UKBB to measure correlations between asthma and additional diseases and traits. Applying LDSC to the UKBB EUR GWAS of 1,008 significantly heritable (heritability Z score > 4) phenotypes and the GBMI leave-UKBB-out meta-analysis of asthma, we obtained pairwise genetic correlation estimates between these phenotypes and asthma. We observed strong correlations (|*r*_*g*_| > 0.4) with 95 of these phenotypes, which spanned prescriptions, PheCodes, and other categories (**Supplementary Table 12**). Digestive system disorders, including gastritis and gastroesophageal reflux disease (GERD), emerged as a disease category with significant and strong genetic correlations with asthma. Although the association between asthma and digestive disorders has not been as well studied, this does reinforce a previous finding of shared genetics between asthma and diseases of the digestive system^9^, indicating that the commonly-observed co-presentation of asthma and gastroesophageal disease in the clinic may be partially due to pleiotropic genetic effects. Our results also showed moderate and significant correlations (*r*_*g*_ ranging from 0.2-0.3) between asthma and neuropsychiatric diseases, like anxiety and depression, and obesity-related traits, like body mass index, which is similarly consistent with previous findings^20,22^.

Leveraging data from another biobank, BBJ, we computed genetic correlation estimates between the GBMI leave-BBJ-out meta-analysis of asthma and 19 significantly heritable disease endpoints in BBJ (**Supplementary Table 13**). COPD showed the strongest and most significant correlation with asthma (*r*_*g*_ = 0.29, p = 6.41×10^−6^), but the notably lower estimate compared to the estimate from the UKBB correlation analyses may be due to differences in phenotype definition and curation. Pollinosis, also known as allergic rhinitis or hay fever, showed moderate correlation with asthma (*r*_*g*_ = 0.28, p = 0.0004), consistent with the correlation results from UKBB (*r*_*g*_ = 0.39, p = 4.60×10^−3^). Comparing the phenotypes with significant SNP heritability estimates in both BBJ and UKBB (**Supplementary Fig. 12**), we found that only COPD has significant genetic correlations with asthma across the biobanks. The rheumatoid arthritis (RA) and type 2 diabetes (T2D) GWAS from UKBB have moderate and significant correlations with asthma, which are partially recapitulated in the BBJ results that showed a moderate but not significant correlation between the BBJ GWAS of RA and of asthma, and a small but significant correlation between the BBJ GWAS of T2D and the GBMI leave-BBJ-out meta-analysis of asthma. Several studies in the literature have reported a relationship between risk for RA and asthma^73–78^, as well as T2D and asthma^79–81^, but more genetic studies in different populations and biobanks are needed to investigate the potential shared genetic architecture of these diseases. Importantly, causal relationships between asthma and genetically correlated phenotypes are not yet well-understood, and other methods such as Mendelian randomization could be applied to identify potential causal associations^82^.

## Discussion

Assembling the largest and most diverse collection of asthma cohorts to date, we conducted a GWAS meta-analysis of 18 biobanks around the world and identified 49 novel associations among a total of 179. Despite the substantial sample sizes of previous meta-analyses of asthma^9^, our results indicated that the heterogeneity and complexity of asthma, like other common, polygenic diseases, will benefit from even larger sample sizes for genomic discovery. We interrogated the overall consistency of genetic effects across the cohorts and found that despite variability in recruitment, continent, sampling strategy, health system design, and disease prevalence, the effects of the loci discovered in the meta-analysis were by and large concordant across the biobanks. Additionally, genetic correlation estimates across ancestries, which ranged from 0.65 to 0.99 for the well-powered ancestry groups, strongly supported the finding that the genetic architecture of asthma is largely shared across the ancestry groups studied.

Importantly, however, the addition of GWAS from more diverse populations aided the discovery of genetic loci with higher frequencies in non-EUR populations that did not reach genome-wide significance in the meta-analysis with only EUR cohorts, highlighting the importance of diversifying genomic studies of asthma. Given the current disproportionate representation of European ancestries, we expect that as the availability of non-EUR GWAS of asthma and other asthma-related diseases and traits continues to increase, it is likely that greater numbers of such variants associated with asthma will be discovered. Previous studies of asthma-related diseases, such as atopic dermatitis, in non-EUR populations have similarly identified additional risk variants that are higher in frequency in other populations but also found highly shared polygenic architecture between populations, mirroring our findings for asthma^83,84^. This study also provides further evidence for substantial genetic overlap between childhood and adult-onset asthma, as well as between asthma and well-known, immune-related comorbidities like COPD and allergic diseases. Additionally, we identified genetic correlations between asthma and less well-studied comorbidities like digestive system disorders, while highlighting additional complexity in the etiology and comorbidities of asthma. For example, gene set enrichment analyses using MAGMA did not yield many shared pathways for asthma and COPD despite the strong genetic correlation.

We also demonstrated that the greater diversity of GBMI improved polygenic prediction in asthma, particularly for populations of non-European ancestry. Previous studies on asthma PRS in the literature have primarily focused on using PRS to predict asthma in pediatric cohorts, and overall found limited performance of PRS^28–30,85^. Most of these studies used the P+T approach, while a recently published paper, Namjou et al. (2022)^32^, applied PRS-CS to the TAGC multi-ancestry GWAS and found improved discriminatory power of their PRS (receiver-operating characteristic area under the curve, or AUC, of 0.66-0.70 across two pediatric cohorts) compared to the prior studies that used P+T. Sordillo et al. (2021)^31^ applied another genome-wide approach, lassosum, to the TAGC data, but their PRS evaluated in adult cohorts showed moderate performance (AUC of 0.51-0.57 across cohorts of different ancestries). While we did not assess the lassosum method, we have shown that the greater sample size and diversity of GBMI compared to TAGC contribute to better performing PRS (**Fig. 5**). However, we also found that differences in prediction power between Bayesian PRS construction methods PRS-CSx and PRS-CS were minimal. This may be due to imbalances in the sample sizes of the discovery cohorts, which may need to be taken into careful consideration when using these methods. Previous studies have found that imbalanced sample sizes across ancestries contribute somewhat unpredictably to varying prediction performances, with a low signal-to-noise ratio in ancestry-matched target populations reducing prediction performance^86^. Therefore, further investigation is needed to fully understand the interplay between sample size and ancestry in the context of polygenic prediction. Ultimately, these analyses highlight the pressing need for more well-powered and ancestrally-diverse resources that will help reduce these imbalances.

We have highlighted the harmonization of the phenotype definitions across biobanks, but it is important to acknowledge that the criteria used, which allowed for both self-reported and PheCode information, are vulnerable to imprecision and variability in the data collected. Self-reported data for asthma is particularly susceptible to imprecision, since it relies on personal recollection of asthma diagnoses that are often given in childhood. On the other hand, PheCodes, which are based on ICD codes, may fail to capture diagnoses made earlier in the lifetime of individuals in hospital-based cohorts. Therefore, including both self-reported and PheCode data -- an approach adopted by some but not all biobanks -- may be optimal for association analyses for asthma. We were limited in our ability to evaluate the effects of phenotype definition on effect size estimation, since only three biobanks used self-reported data, and 2 of the 3 biobanks (TWB and BBJ) only have participants of EAS ancestry. However, we compared the asthma GWAS derived from self-reported vs. PheCode data in UKBB and found high genetic correlation (*r*_*g*_ (se) *=* 0.95 (0.01)) between the GWAS. This provides some evidence that variation in phenotype definition may not significantly influence genetic discovery, but we cannot confirm the same pattern for all biobanks in GBMI and especially for other diseases. However, given the relative alignment of genetic effects across the biobanks, we would expect that minor differences in phenotype definition would not substantially change the association results for asthma.

Additionally, we acknowledge that since the definitions used here for asthma and COPD do not exclude individuals with concurrent diagnoses, we are not able to fully distinguish the distinct biological pathways affecting asthma and COPD. Comorbidity rates of asthma and COPD reported in the literature range across studies but population-based estimates generally are low, around 2-3%^87,88^, while hospital-based prevalence estimates tend to be higher, around 13%^89^. Among biobanks participating in GBMI, for example, 15.5% of all individuals with asthma in UKBB have a concurrent COPD diagnosis, 21% in BioVU, and 7.4% in BBJ. A previous study found that using stricter definitions of asthma, such as excluding subjects with COPD, resulted in stronger association signals for some of the asthma-associated loci^7^. However, it is important to note that if we excluded participants with a COPD diagnosis, we would not have a fully representative sample of the participants in GBMI with asthma. As has been documented in other studies^90,91^, this could induce selection bias, or collider bias, which could lead to biased genetic associations. Most of the previous genetic studies of asthma in the literature did not exclude individuals with COPD from analyses. However, in the COA and AOA analyses, we do exclude participants with a COPD diagnosis to avoid confounding from potential misclassifications of adult-onset asthma and COPD. We also note that estimates of genetic correlation by LDSC are not biased by sample overlap^92^. In fact, this has been explored in the context of asthma and allergic diseases, where *r*_*g*_ estimates from LDSC were shown to be robust to overlapping cases and/or controls^21^.

We also recognize the importance of analyzing environmental factors in conjunction with genetic factors for a disease that is heavily influenced by the environment. Our genetic analyses offer insight into the potential shared biological pathways that may be differentially affected by non-genetic factors, but we were not able to explicitly investigate environmental effects given the lack of available environmental exposure data among the biobanks. The high degree of alignment among genetic associations, coupled with the large variability in asthma prevalence, points to a particularly important role of the environment for asthma risk across populations. Gaining a greater understanding of the specific non-genetic factors that contribute to asthma development in different environments may help guide more accurate disease prediction across populations.

This study, and importantly the data sharing across biobanks facilitated by this initiative, have laid the groundwork for deeper dives into the shared and distinct genetic signatures of asthma subtypes. We were able to stratify two participating biobanks, UKBB and FinnGen, into COA and AOA based on the participants’ ages at first diagnosis. While we found that the GBMI asthma meta-analysis of all biobanks containing both subtypes identified many of the loci contributing to these subtypes, the age-of-onset-stratified meta-analyses uncovered additional subtype-specific loci. Of the top loci associated with COA and AOA, 11 and 12 loci, respectively, (1) did not overlap with a top locus in the other subgroup meta-analysis; and (2) were evaluated in the all-asthma GBMI meta-analysis (i.e. in more than 3 GBMI biobanks) but did not reach genome-wide significance in the meta-analysis (**Supplementary Table 11**). Due to the limited availability of age at first diagnosis information across the biobanks, we were not able to explore age-dependent associations further, but with sufficient scale, it is likely that more of the distinct genetic architectures of COA and AOA will be uncovered.

As the examples from this study demonstrate, with broader sharing of more extensive phenotype data, biobanks are well-positioned to facilitate not only general locus discovery, but also advance the study of disease subtypes and comorbidities. The inclusion at a continuously increasing scale of individuals of diverse ancestries will accelerate novel variant and gene discovery. This will more quickly expand the set of genetic findings from which biological inference can be drawn, as well as ensure that predictive models derived from genetic and environmental risk factors will be as accurate and informative for individuals of all ancestries and geographical locations as possible.

## Supporting information

GBMI Banner Author List

## Data Availability

The all-biobank GWAS summary statistics are publicly available for downloading at https://www.globalbiobankmeta.org/resources and can be browsed at the PheWeb Browser (http://results.globalbiobankmeta.org).

## Acknowledgements

We acknowledge helpful comments from Cristen Willer, Hailiang Huang, Yunfeng Ruan, Tian Ge and Chris Gignoux. A.R.M is funded by the K99/R00MH117229. S.N. is supported by Takeda Science Foundation. Y.O. is supported by JSPS KAKENHI (22H00476), and AMED (JP21gm4010006, JP22km0405211, JP22ek0410075, JP22km0405217, JP22ek0109594), JST Moonshot R&D (JPMJMS2021, JPMJMS2024), Takeda Science Foundation, and Bioinformatics Initiative of Osaka University Graduate School of Medicine, Osaka University. R.G. is supported by the T32AG000222.

## Author Contributions

Study design: K.T., A.R.M., M.J.D., B.M.N.

Data collection/contribution: W.Z., Y.O., T.M.

Data analysis: K.T., W.Z., Y.W., M.K., S.N., R.G., L.M., L.N.

Writing: K.T., A.R.M., S.N., R.G.

Revision: K.T., A.R.M., Y.W., R.G., M.J.D.

## Declaration of Interests

M.J.D. is a founder of Maze Therapeutics. B.M.N. is a member of the scientific advisory board at Deep Genomics and consultant for Camp4 Therapeutics, Takeda Pharmaceutical, and Biogen.

## STAR Methods

### Asthma phenotype definitions for association analyses

The phenotype definition guidelines that were developed by GBMI and shared with all participating biobanks can be found in Zhou et al. (2021)^33^. Disease endpoints, including asthma, were defined following the PheCode maps, which maps ICD-9 or ICD-10 codes to PheCodes^93^. Asthma cases were all study participants with the asthma PheCode, and controls were all study participants without the asthma PheCode (or asthma-related PheCodes). Biobanks that did not have ICD codes primarily used self-reported data (**Supplementary Table 3**).

### Principal components (PC) projection for genetic ancestry comparison

To compare the genetic ancestries represented in different biobanks, we used pre-computed loadings of genetic markers shared across all biobanks and the reference data containing 1000 Genomes (1000G) and the Human Genome Diversity Project (HGDP) to project biobank participants to the same principal components space. 179,195 genetic variants were genotyped/imputed in all biobanks, among which 168,899 are also in the 1000 Genomes^34^ and HGDP^94^. The weights corresponding to principal components for those markers were estimated based on the PCA analysis for the reference samples with known ancestry in 1000G and HGDP and shared among biobanks. Biobanks then generated PC loadings based on the pre-estimated weights of those markers. More details are described in Zhou et al. (2021)^33^.

### Meta-analysis for asthma in GBMI

We performed fixed-effects meta-analysis with inverse variance weighting for 18 biobanks in GBMI: China Kadoorie Biobank (CKB), Generation Scotland (GS), Lifelines, QSKIN, East London Genes & Health (GNH), HUNT, UCLA Precision Health Biobank (UCLA), Colorado Center for Personalized Medicine (CCPM), Mass General Brigham (MGB), BioVU, BioMe, Michigan Genomics Initiative (MGI), BioBank Japan (BBJ), Estonian Biobank (ESTBB), deCODE Genetics (DECODE), FinnGen, Taiwan Biobank (TWB), and UK Biobank (UKBB). Basic information on the biobanks are described in Zhou et al. (2021)^33^, as well as details on the genotyping, imputation, GWAS, post-GWAS quality control, and meta-analysis procedures^33^. In brief, genetic variants with minor allele count (MAC) < 20 and imputation score < 0.3 were excluded from the analyses. Genetic variants with different allele frequencies (AF) compared to gnomAD^95^ (Mahalanobis distance between AF-GWAS and AF-gnomAD > 3 standard deviations away from the mean) were also excluded. Altogether, these cohorts had a total sample size of 153,763 cases and 1,647,022 controls (**Supplementary Table 1**). GWAS meta-analyses were first conducted within continental ancestry groups to control for population stratification. 5,051 cases and 27,607 controls were of African (AFR) ancestry; 4,069 cases and 14,104 controls were of Admixed American (AMR) ancestry; 18,549 cases and 322,655 controls were of East Asian (EAS) ancestry; 121,940 cases and 1,254,131 were of European (EUR) ancestry; 139 cases and 1,434 controls were of Middle Eastern (MID) ancestry; and 4,015 cases and 27,091 controls were of Central and South Asian (CSA) ancestry.

We also performed fixed-effects meta-analysis with inverse variance weighting for 4 additional biobanks that served as independent replication studies: Canadian Partnership for Tomorrow’s Health (CanPath), Qatar Biobank (QBB), Biobank of the Americas (BBofA), and Penn Medicine Biobank (PMBB). Collectively, these cohorts had a total sample size of 9,991 cases and 63,605 controls (**Supplementary Table 1**). More information on these biobanks are also described in Zhou et al. (2021)^33^.

### Index variant and locus definitions

We used a threshold of p < 5×10^−8^ to identify SNPs with a genome-wide significant effect. To identify loci, we used a window size of 500 kb upstream and downstream of the SNPs with the strongest evidence of association in the meta-analysis, and merged overlapping regions until no genome-wide significant variants were detected within the ± 500 kb region. To designate loci as previously discovered or potentially novel, we compiled a list of known asthma-associated SNPs (p < 5×10^−8^) from the associations collected by El-Husseini et al. (2020)^6^ and listed in the GWAS catalog (as of 11/14/2021)^96^. We extended 500 kb upstream and downstream of each of these variants to define a locus, and intersected these regions with the loci defined by the index variants in our meta-analysis to identify any overlaps. We annotated genetic variants with the nearest genes using ANNOVAR^97^ and putative loss-of-function using VEP^98^ with the LOFTEE plug^95^ as implemented in Hail^33^. We also annotated whether the index or tagging variants (r^2^ > 0.8) of asthma were fine-mapped in any of the cis-eQTL fine-mapping resources. We retrieved cis-eQTL fine-mapped variants with posterior inclusion probability (PIP) > 0.9 in any tissues and cell types from the GTEx v8^99^ and eQTL catalogue release 4^100^. Fine-mapping was conducted using SuSiE^101^ with summary statistics and covariate-adjusted in-sample LD matrix as described previously^102,103^.

### Index SNP effects across biobanks

To estimate the correlation of SNP effects for the 179 top loci between one specific biobank and the leave-that-biobank-out meta-analysis, we used the method proposed by Qi et al. (2018) using GWAS summary statistics^50^ (**Supplementary Table 5**). Specifically, the method directly calculates SNP effect correlation as:

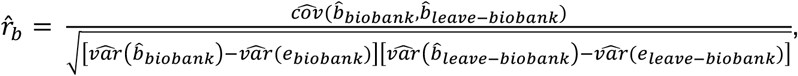

where 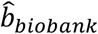 and 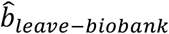 denote the estimated SNP effects from GWAS conducted in one specific biobank and from GWAS performed in the leave-that-biobank-out meta-analysis, respectively. The 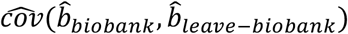 is calculated as the sampling covariance between 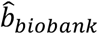 and 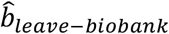The 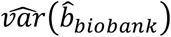 and 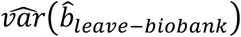 are the estimated variances of 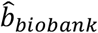 and 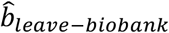 separately. The 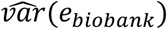 and 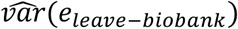 are the estimated variance of the estimation errors of 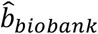 and 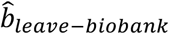 which are approximated as the mean of the squared standard errors of estimated SNP effect (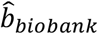 and 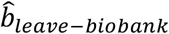) across all the top-associated SNPs, respectively. The standard error of 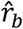 is obtained through the jackknife approach by leaving one SNP out each time. SNPs with large standard errors in CKB and HUNT (chr12:123241280:T:C and chr17:7878812:T:C, respectively) were excluded from these analyses.

Then, for the index variants present in each biobank, we computed:

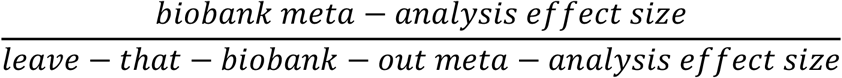

for the biobank and leave-that-biobank-out pair. We took the average ratio across the index variants for each biobank and leave-that-biobank-out pair. We then used the regression method introduced in Deming et al. (1943), which considers the errors in both the *X*- and *Y*-variables, to compare the effect sizes of these SNPs in each biobank GWAS with their effects in the leave-that-biobank-out meta-analysis^35^. We set the intercept equal to 0 for these analyses.

### Ancestry-specific heterogeneity

To assess heterogeneity of per-SNP effect sizes for the 179 top loci across ancestries in GBMI, we conducted ancestry-specific meta-analyses of the five most well-powered ancestry groups in GBMI (EUR, AFR, AMR, EAS, and CSA). We applied the Cochran’s Q test^104^ to the SNP effects in the ancestry-specific meta-analyses and identified SNPs with significant heterogeneity based on a Bonferroni-corrected p-value cut-off of 0.05/169 = 0.0003, accounting for the number of SNPs present in all studies (**Supplementary Table 7**). Regions displaying heterogeneity in effects across ancestry groups were visualized using the LocalZoom tool^105^.

### Polygenic risk scores

A description of the PRS analyses conducted using PRS-CS^57^, as well as the leave-one-biobank-out meta-analysis strategy applied, is provided in Wang et al. (2021)^56^.

We used PRS-CSx, which jointly models GWAS summary statistics from populations of different ancestries and returns posterior SNP effect size estimates for each input population^58^. We applied this method to the AMR, AFR, CSA, EAS, and EUR ancestry-specific meta-analyses, which served as the discovery data for PRS construction. For the ancestry-specific meta-analysis that matched the ancestry of the target cohort, we excluded the target cohort. We evaluated the predictive performance of the PRS in 4 target cohorts: 1) AFR ancestry individuals in UKBB (849 cases, 5190 controls), 2) CSA ancestry individuals in UKBB (1232 cases, 6744 controls), 3) EAS ancestry individuals in BBJ that were part of a randomly-selected 1k holdout set (500 cases, 500 controls), and 4) EUR ancestry individuals in UKBB that were part of a randomly-selected 10k holdout set (1164 cases, 7577 controls). We also evaluated the PRS in an additional randomly-selected 1k holdout set (131 cases, 869 controls) of EUR ancestry individuals in UKBB. As an example, for the AFR ancestry individuals, the full set of discovery data for PRS construction consisted of the AMR, CSA, EAS, and EUR ancestry-specific meta-analyses, as well as the AFR ancestry-specific meta-analysis excluding the AFR ancestry individuals in UKBB. The same strategy was applied to the other 3 target cohorts (**Supplementary Table 9**). We used ancestry-matched LD reference panels from UKBB data and the default PRS-CSx settings for all input parameters. We evenly and randomly split cases and controls in the target cohorts into validation and testing subsets. Using the posterior SNP effect size estimates from PRS-CSx, we computed one PRS per discovery population for the validation subsets to learn the optimal linear combination of the ancestry-specific PRS using PLINK v1.9^106,107^. Then, with these weights, we evaluated the prediction accuracy of this linear combination of PRS in the testing subset. We reported the average prediction accuracy, measured by variance explained on the liability scale 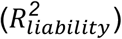, estimated using the prevalence of asthma in the BBJ biobank for the EAS target cohort and in the UKBB biobank for the other target cohorts, across 100 random splits.

### Asthma age-of-onset subtype GWAS and meta-analyses

#### UKBB

We first identified EUR individuals in UKBB with an asthma diagnosis based on information from the asthma PheCode or field 20002, which has self-reported diagnoses from verbal interviews. We then excluded individuals with either (1) a COPD diagnosis based on the COPD PheCode or field 20002, (2) missing information for field 3786, which has age at first asthma diagnosis information, (3) an asthma diagnosis after age 60 based on field 3786, or (4) greater than 10 years between the age reported in field 3786 and the age reported in field 22147, another age at first asthma diagnosis field that only a subset of participants filled out as part of a follow-up questionnaire. Then, using the age at first diagnosis reported in field 3786, we divided these individuals into asthma age of onset groups: those with diagnoses at or before age 19 were childhood-onset (n = 12,577) and after age 19 were adult-onset (n = 23,533). We then conducted separate COA and AOA GWAS using Scalable and Accurate Implementation of GEneralized mixed model (SAIGE)^108^. The same set of controls was used (n = 359,116) for both GWAS, derived based on the PheCode guidelines provided by GBMI^33^.

#### FinnGen

We identified individuals with an asthma diagnosis based on the PheCode guidelines provided by GBMI^33^ (**Supplementary Table 3**). We excluded individuals with either (1) a COPD diagnosis based on the COPD PheCode definition, or (2) an asthma diagnosis after age 60. Those with diagnoses at or before age 19 were childhood-onset (n = 8,387) and after age 19 were adult-onset (n = 33,191). We conducted separate COA and AOA GWAS using SAIGE^108^. The same set of controls was used (n = 314,898) for both GWAS, derived based on the PheCode guidelines provided by GBMI^33^.

#### Meta-analyses

We performed fixed-effects meta-analysis with inverse variance weighting for the COA GWAS from UKBB and FinnGen and the AOA GWAS from both biobanks. We used linkage-disequilibrium score correlation (LDSC)^92^ to compute pairwise genetic correlations (*r*_*g*_) between (1) the subtype meta-analyses, (2) each subtype meta-analysis and the GBMI COPD meta-analysis, (3) each subtype GWAS from UKBB and the GBMI all-asthma leave-UKBB-out meta-analysis, and (4) each subtype GWAS from FinnGen and the GBMI all-asthma leave-FinnGen-out meta-analysis. Finally, using the regression method introduced in Deming et al. (1943)^35^, we compared the effect sizes of the 179 index variants discovered in the GBMI all-asthma meta-analysis in each subtype meta-analysis. We set the intercept equal to 0 for this analysis.

### Genetic correlation in UKBB and BBJ

Using LDSC, we estimated *r*_*g*_ between all EUR-ancestry UKBB phenotypes with heritability Z-score > 4 and (1) the GBMI leave-UKBB-out meta-analysis for asthma and (2) the UKBB EUR-ancestry GWAS of asthma (PheCode ID 495 in UKBB). The heritability Z-scores were obtained from the stratified-LDSC (S-LDSC) computations of heritability reported by the Pan-UK Biobank team^109–111^. Summary statistics from the UKBB EUR GWAS were obtained from the Pan-UK Biobank team as well^111^.

We also used LDSC^92^ to compute *r*_*g*_ between 48 phenotypes in BioBank Japan (BBJ) and (1) the GBMI leave-BBJ-out meta-analysis for asthma and (2) the BBJ GWAS of asthma. We used publicly available GWAS summary statistics for all traits^112–114^. Genetic correlation results were visualized using the R corrplot package^115^.

### Gene- and pathway-based enrichment for asthma and COPD

Fixed-effects meta-analysis with inverse variance weighting was also performed for 16 biobanks in GBMI with COPD data: BBJ, BioMe, BioVU, CCPM, CKB, ESTBB, FinnGen, GNH, GS, HUNT, Lifelines, MGB, MGI, TWB, UCLA, and UKBB. The same processing and methods were applied here as for the asthma meta-analysis. These cohorts had a total sample size of 81,568 cases and 1,310,798 controls. COPD cases were defined based on the COPD PheCode, and controls were all study participants without the COPD or COPD-related PheCodes. Biobanks that did not have ICD codes available used spirometry data (Lifelines) or self-reported data (TWB). Details can be found in Zhou et al. (2021)^33^. Meta-analyses were also conducted within continental ancestry groups: 19,044 cases and 310,689 controls of EAS ancestry, 1,978 cases and 27,704 controls of AFR ancestry, and 58,559 cases and 937,358 controls of EUR ancestry.

#### MAGMA

We used Multi-marker Analysis of GenoMic Annotation (MAGMA)^65^ v1.09b for gene prioritization and gene-set enrichment analyses, applying this method to the GBMI asthma EUR, AFR, EAS, and CSA ancestry-specific meta-analyses and the GBMI COPD EUR, AFR, and EAS ancestry-specific meta-analyses. For the gene-level analyses in MAGMA, we first mapped the SNPs to the provided list of genes using a window size of 20kb, and then performed gene analysis using the ancestry-matched 1000G LD reference panels to account for LD structure. Gene-set enrichment was performed using the default settings to correct for gene length, gene density, and the inverse mean minor allele count. The gene sets used were the c2, “curated gene sets,” and c5, “ontology gene sets,” obtained from the Molecular Signatures Database v7.4^68^.

#### DEPICT

We also used Data-driven Expression-Prioritized Integration for Complex Traits (DEPICT)^66^, which performs gene prioritization based on correlation of genes across gene sets. We used a 1000G LD reference panel from individuals of EUR ancestry to calculate LD and identify tag SNPs from GWAS results. We report results from the gene prioritization using a p-value threshold of 5×10^−8^ and a minimum of 10 index variants. We defined significant enrichment results by FDR < 0.05. Full details can be found in Zhou et al. (2021)^33^.

#### PoPS

We used another gene prioritization method, Polygenic Priority Score (PoPS), to identify potential causal genes^67^. PoPS performs gene prioritization based on the integration of GWAS data with gene expression, biological pathway, and predicted protein-protein interaction data. We similarly used a 1000G LD reference panel from individuals of EUR ancestry to obtain gene-level associations. Next, MAGMA was applied to integrate the gene-level associations and gene-gene correlations to perform enrichment analysis for gene features selection. Finally, we computed a PoPS score by fitting a joint model with all the selected features simultaneously. We considered genes with a PoPS score in the top one percentile as the prioritized genes. Full details can be found in Zhou et al. (2021)^33^.

### Resource Availability

#### Data and Code Availability

The all-biobank GWAS summary statistics are publicly available for downloading at https://www.globalbiobankmeta.org/resources and can be browsed at the PheWeb Browser (http://results.globalbiobankmeta.org). Custom scripts used for quality control, meta-analysis, and loci definition are available at https://github.com/globalbiobankmeta. Other analyses utilized publicly available tools: the R deming package for Deming regression^116^, PRS-CSx for polygenic prediction (https://github.com/getian107/PRScsx), LDSC for genetic correlation (https://github.com/bulik/ldsc), and MAGMA v1.09b for gene-set enrichment (https://ctg.cncr.nl/software/magma).

## Supplementary Information

### Supplementary Figures

**Supplementary Figure 1.**
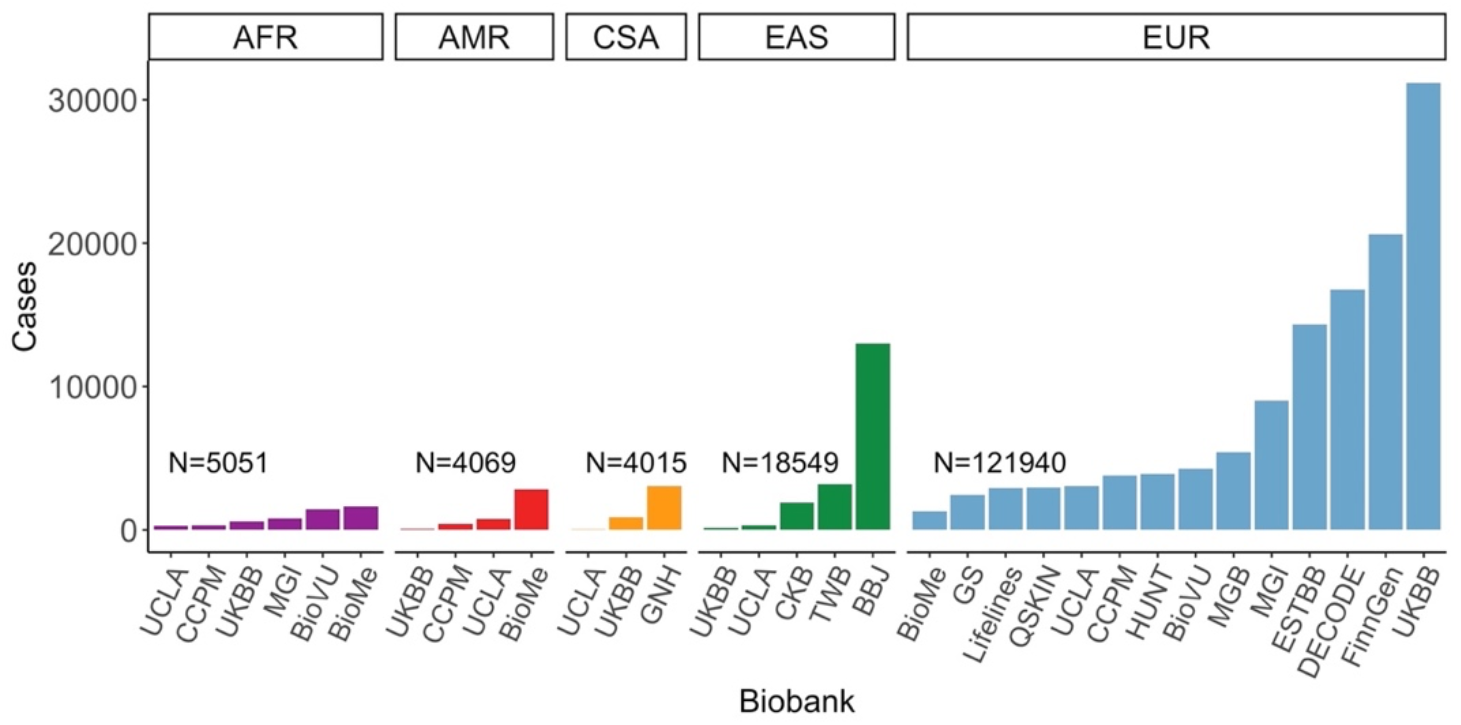
Asthma cases in discovery biobanks stratified by ancestry group. GBMI biobank participants were projected to the same principal components space using pre-computed loadings of genetic markers to compare the genetic ancestries represented in each biobank, indicated on the x-axis. N indicates the total number of cases per ancestry group.

**Supplementary Figure 2.**
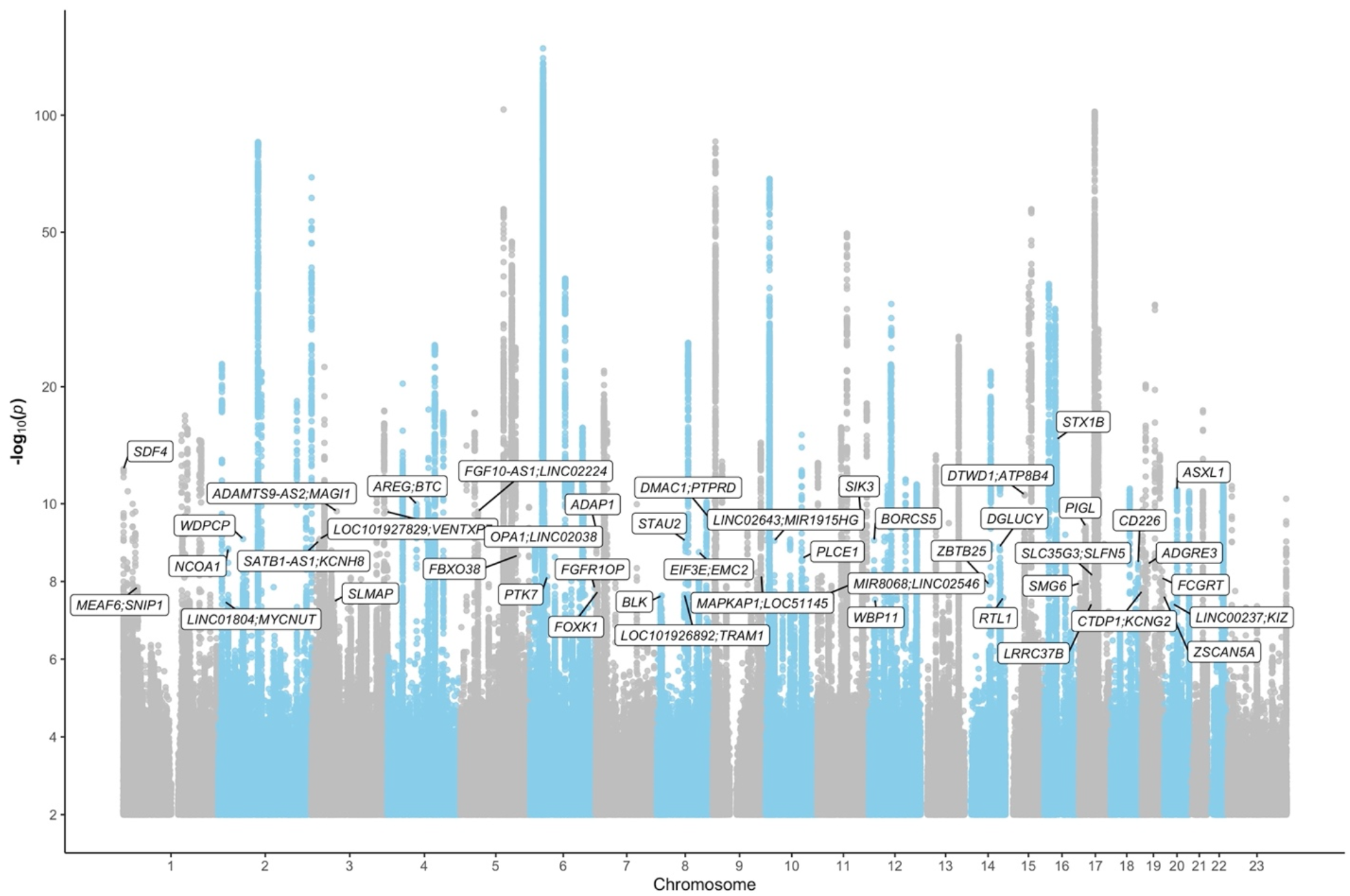
GBMI meta-analysis association results. Nearest genes to the novel loci are highlighted.

**Supplementary Figure 3.**
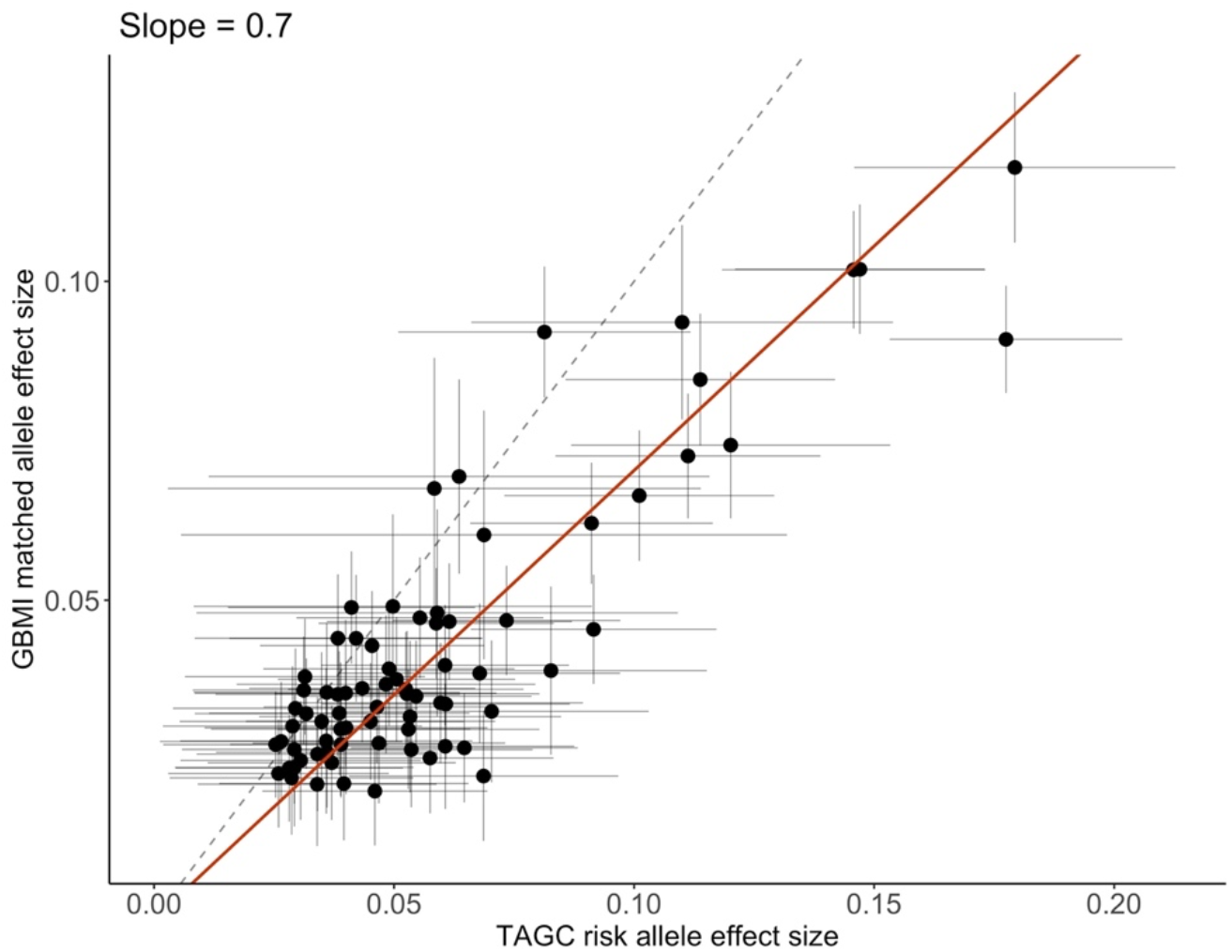
GBMI index variants in TAGC. 76 of the 179 index variants associated with asthma discovered in the GBMI meta-analysis were found in the TAGC meta-analysis of asthma, or had a tagging variant (r2 > 0.8) in the TAGC study, with a p-value < 0.05^9^. The effect sizes of these 76 variants as estimated in the TAGC vs. GBMI meta-analyses were compared using the Deming regression method^35^. The intercept was set to be 0; the slope estimated from the regression analysis is reported here.

**Supplementary Figure 4.**
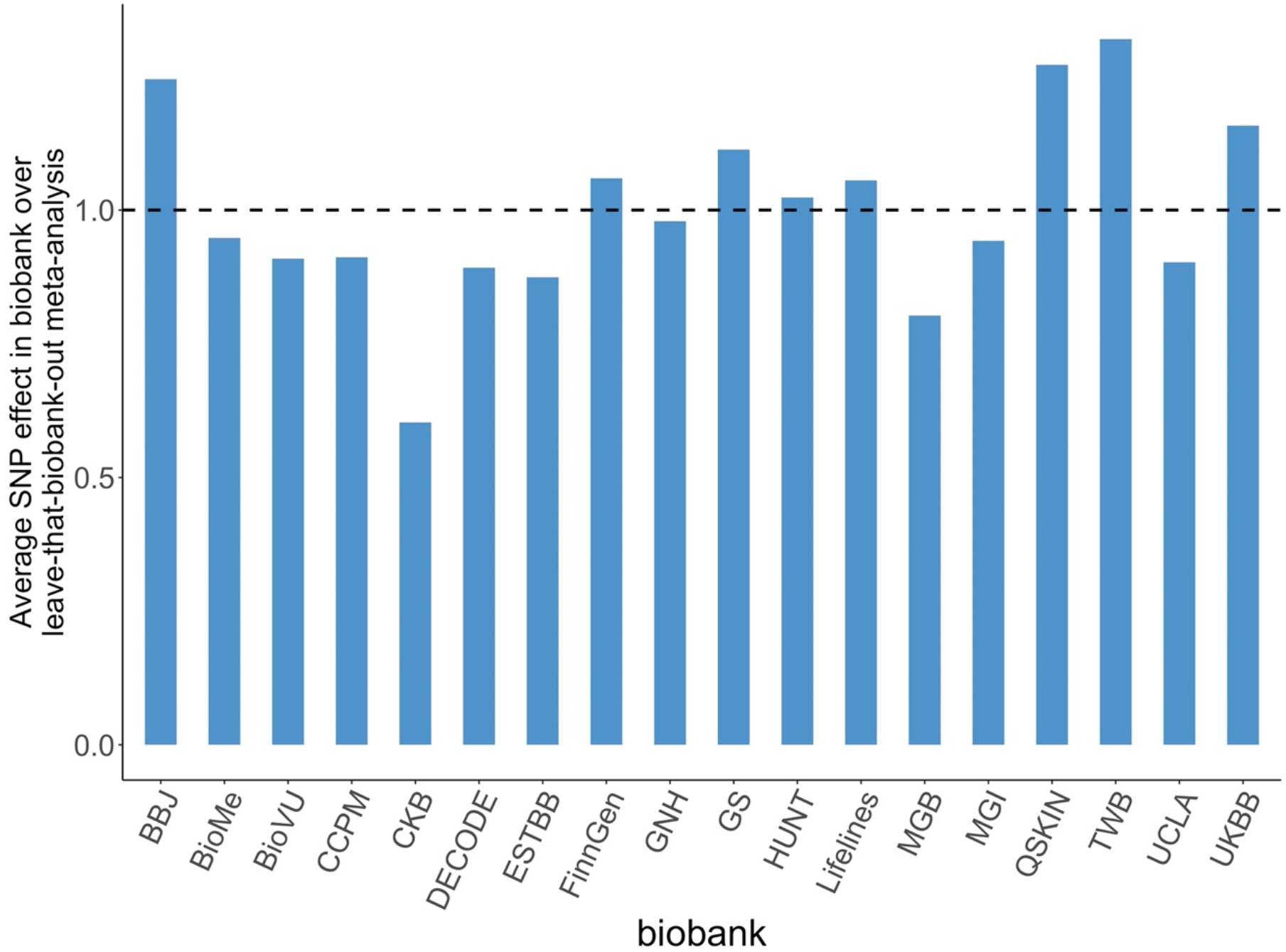
Consistency of asthma index variants across biobanks. For each biobank shown on the x-axis, we computed the average ratio of effect sizes of the index variants in the biobank vs. in the corresponding leave-that-biobank-out meta-analysis.

**Supplementary Figure 5.**
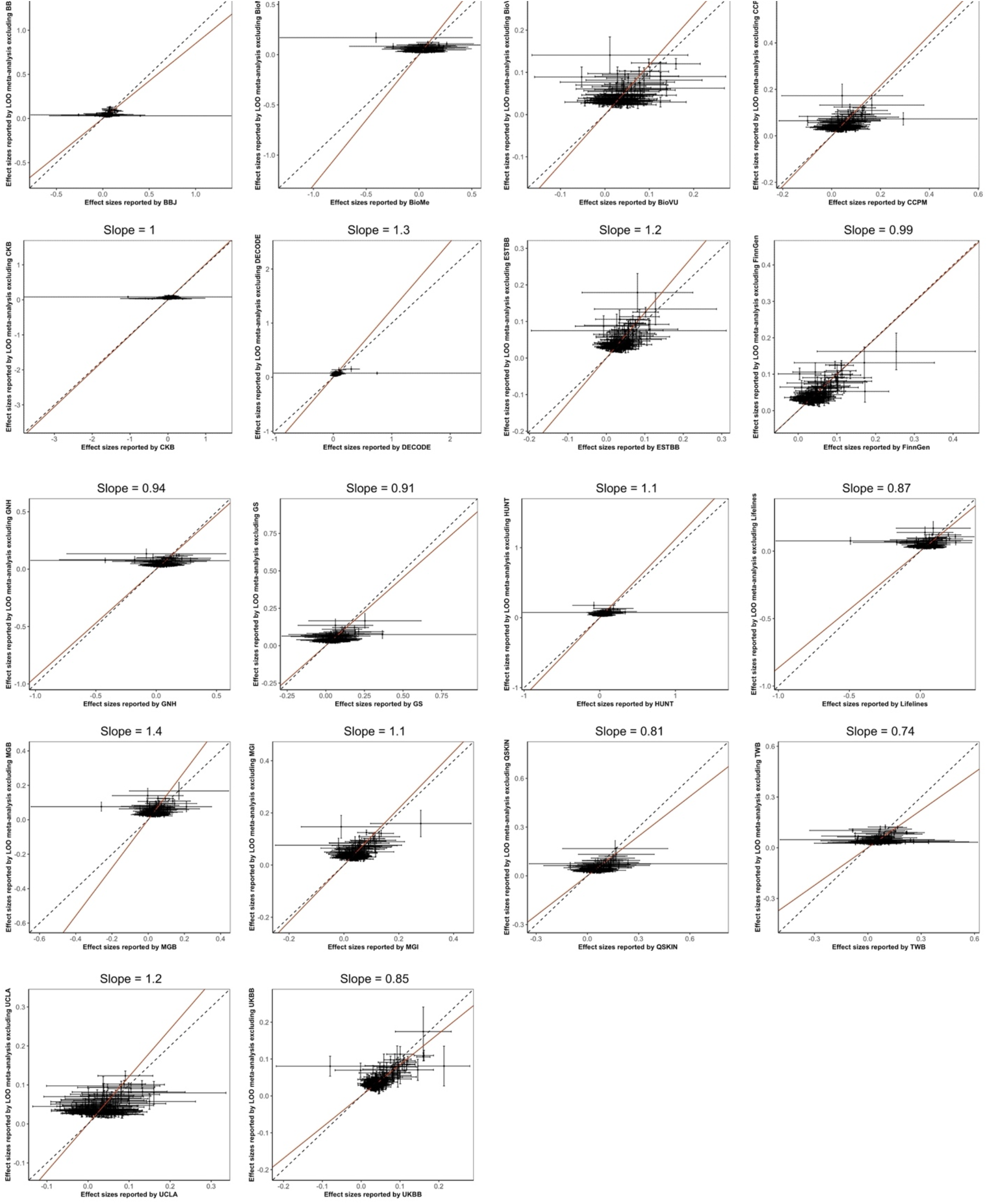
Consistency of asthma index variants across biobanks using Deming regression. The effect sizes of the asthma index variants as estimated in each biobank GWAS vs. in the corresponding leave-that-biobank-out meta-analysis were compared using the Deming regression method^35^. Intercepts were set to be 0; slopes from the regression analyses are reported here.

**Supplementary Figure 6.**
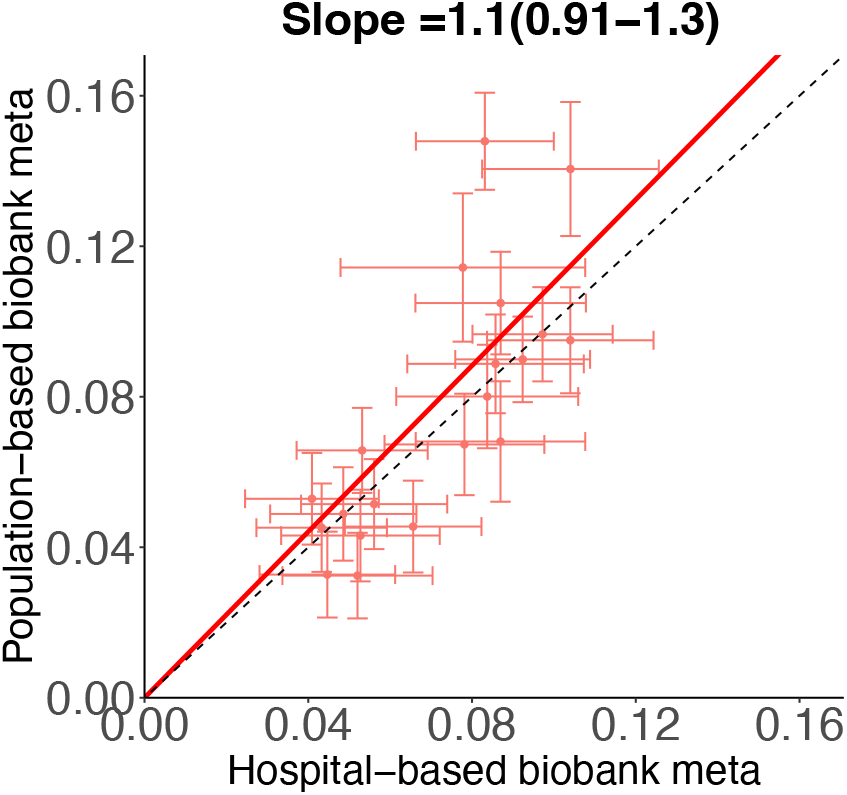
Consistency of asthma index variants across biobanks with different ascertainment. The effect sizes of the asthma index variants as estimated in the meta-analyses of the hospital-vs. population-based biobanks, using the SNPs with p-value < 1×10^−6^ in both meta-analyses, were compared using the Deming regression method^35^. The intercept was set to be 0, and the slope and corresponding 95% confidence interval are reported here.

**Supplementary Figure 7.**
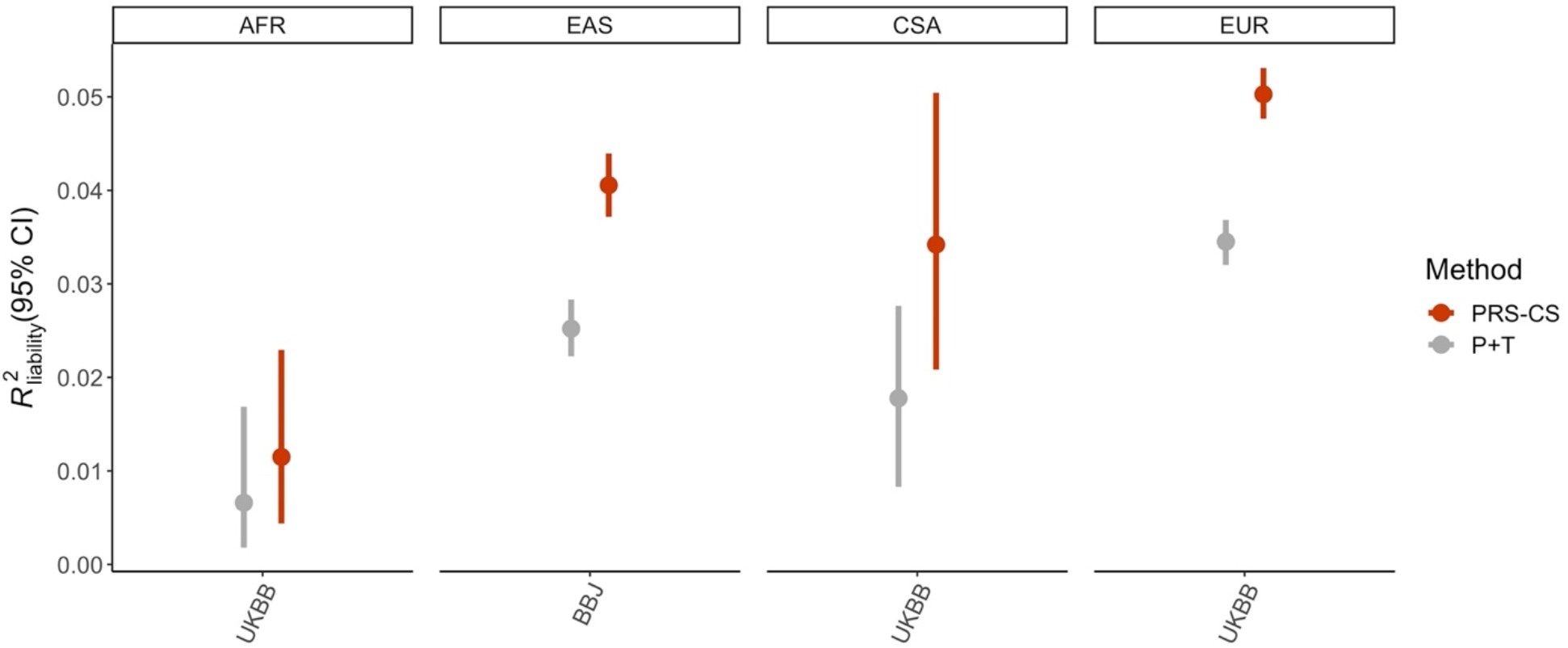
PRS performance of P+T vs. PRS-CS across target cohorts of different ancestries. Each panel represents a target cohort, with the ancestry of the target cohort on the top and the biobank which the target cohort is from on the bottom x-axis. This figure was adapted from Wang et al. (2021)^56^.

**Supplementary Figure 8.**
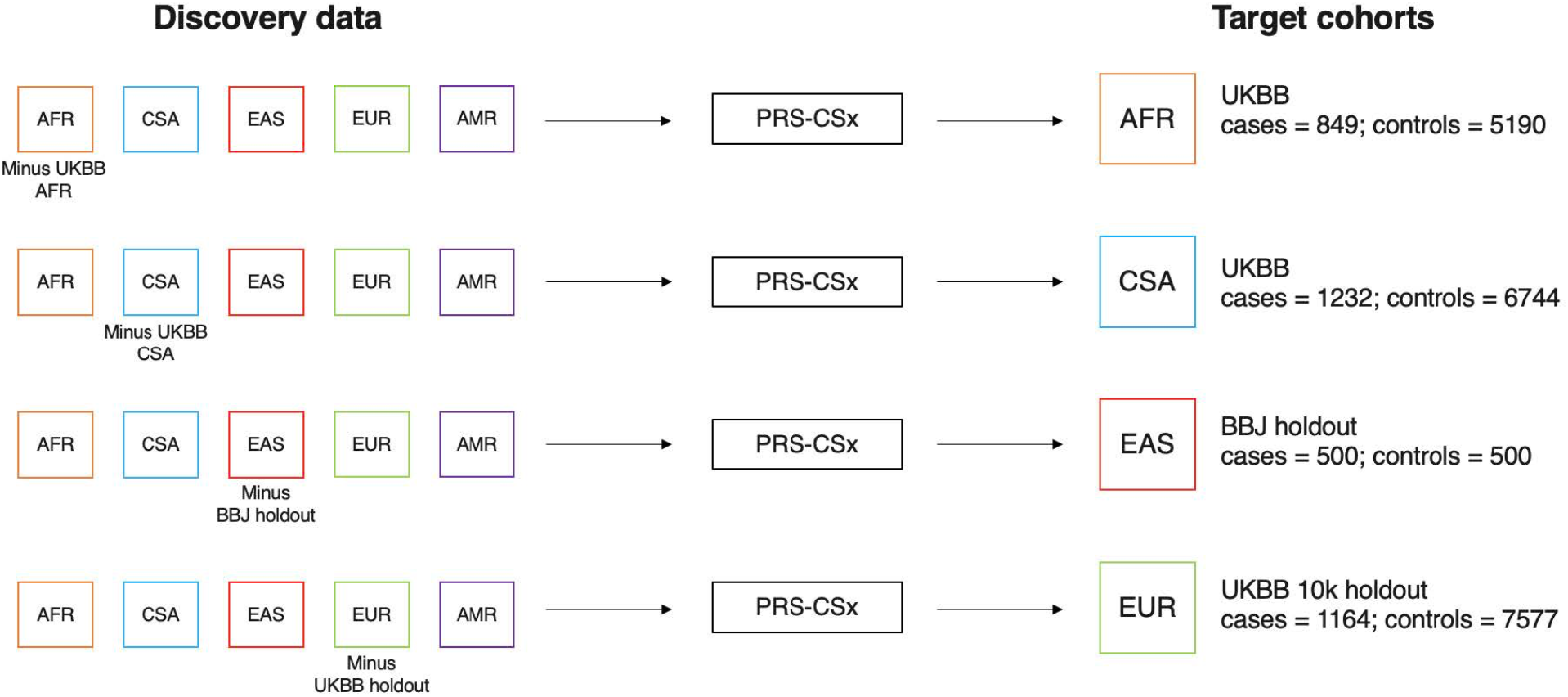
Workflow for PRS-CSx analyses. The discovery data consisted of ancestry-specific meta-analyses, indicated by the squares on the left, that were inputs for PRS-CSx^58^. PRS-CSx returned separate sets of posterior effect size estimates for each input dataset, which were then used to construct PRS. The target cohorts were randomly evenly split; optimal weights for the linear combination of the PRS were selected in one subset and the linear combination of the PRS was evaluated in the other subset.

**Supplementary Figure 9.**
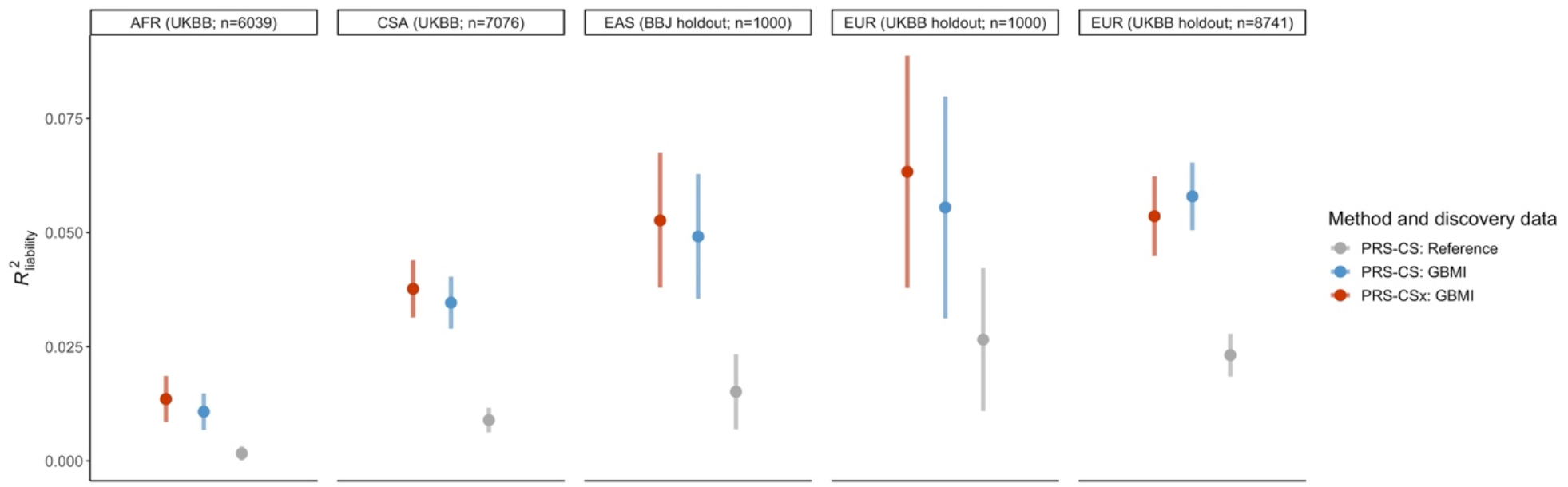
PRS performance in downsampled EUR target cohort. Fig. 5 is extended here to include results from PRS evaluated in a target cohort of 1,000 randomly selected individuals from the EUR UKBB 10k holdout. Discovery datasets and methods used were the same as described in Fig. 5.

**Supplementary Figure 10.**
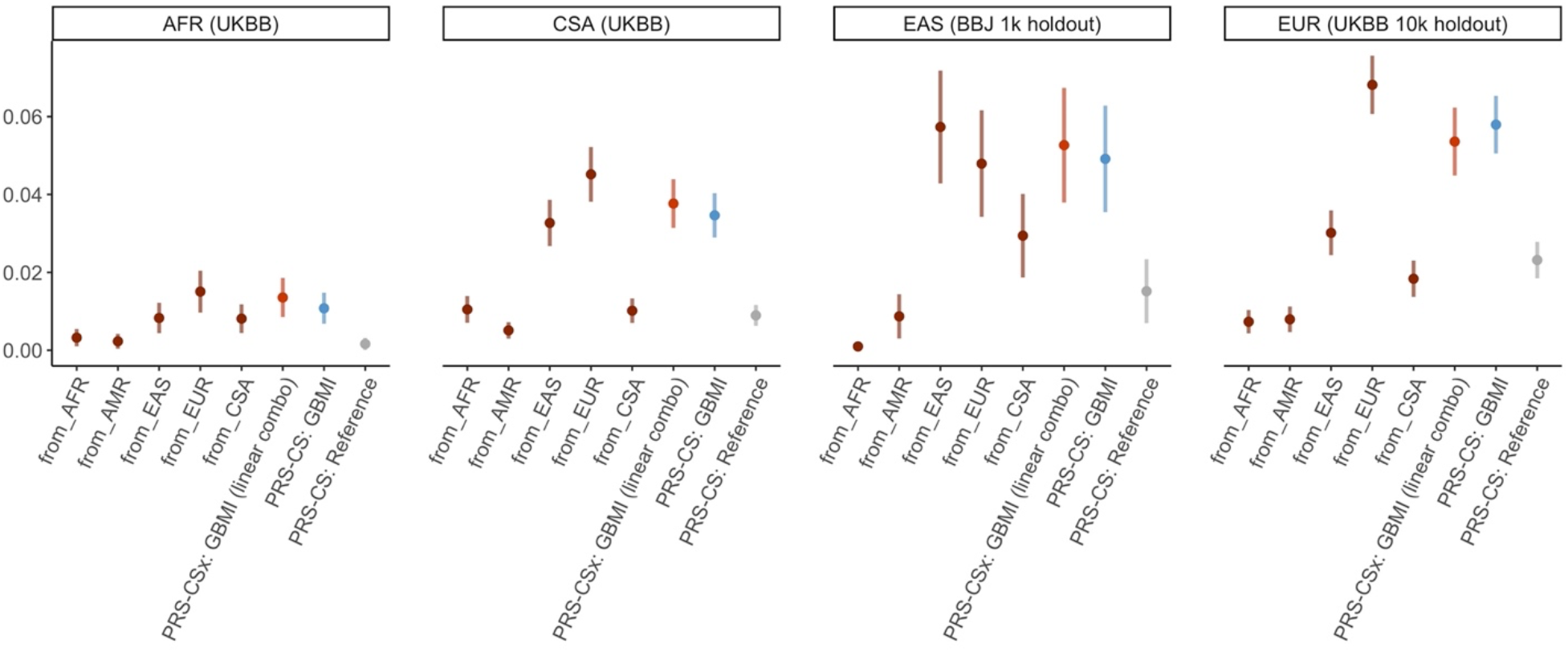
PRS performance of individual PRS vs. linear combination of PRS using PRS-CSx across ancestries. Each panel represents a target cohort. The performance of the individual PRS, computed from a single set of posterior effect size estimates corresponding to each input ancestry population from PRS-CSx, is plotted here. The prediction accuracy of the linear combination of the PRS from PRS-CSx, as well as the PRS from the PRS-CS analyses (shown in Fig. 5), are also plotted for comparison. PRS-CS results used the GBMI leave-BBJ-out meta-analysis and GBMI leave-UKBB-out meta-analysis as discovery data for the BBJ and all UKBB target cohorts, respectively^56^. The reference dataset was the TAGC meta-analysis^9^. Error bars represent standard deviation of R^2^ on the liability scale across 100 replicates.

**Supplementary Figure 11.**
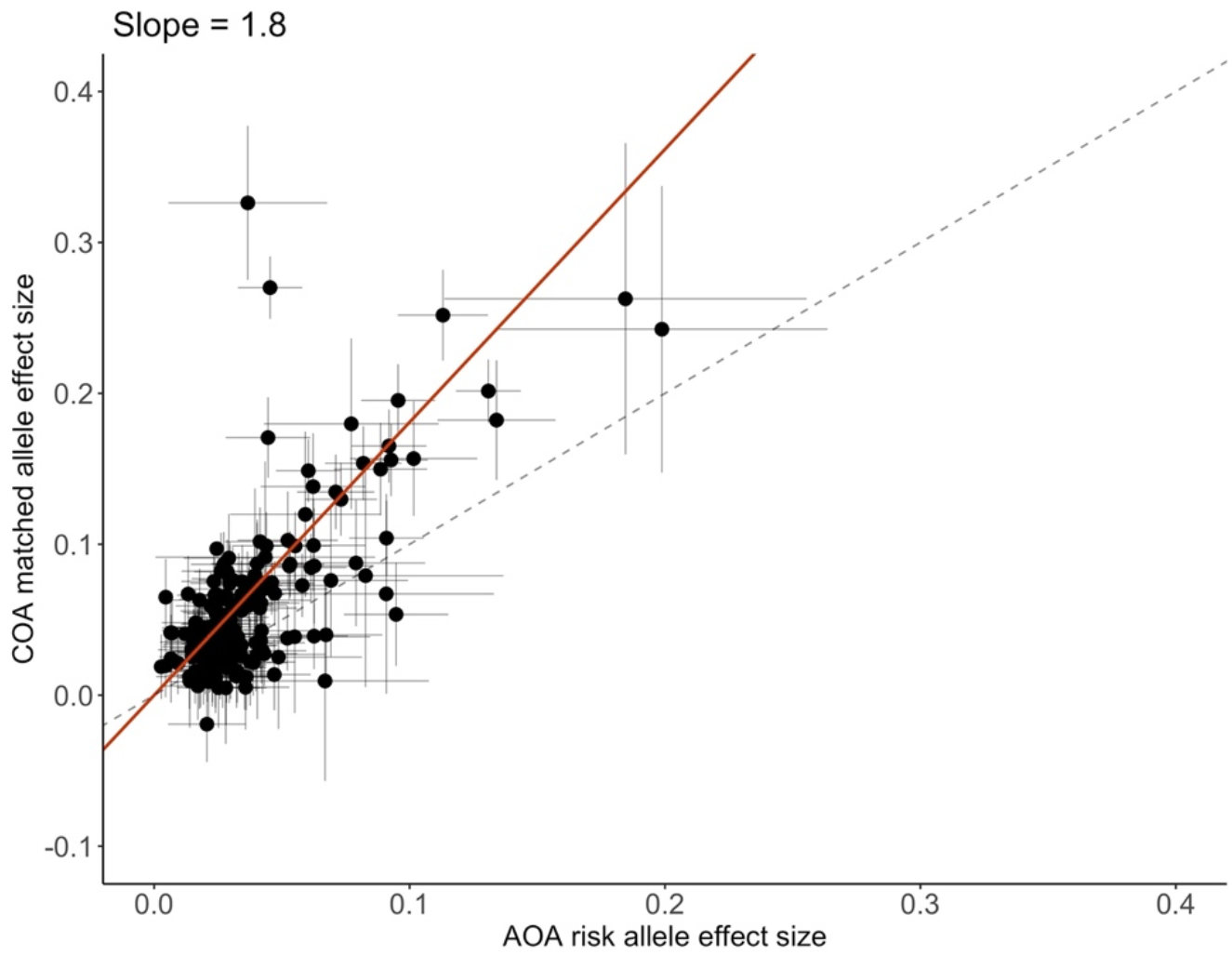
Effect size estimates of asthma index variants in COA vs. AOA meta-analyses. The effect sizes of the 179 index variants discovered in the all-asthma meta-analysis as estimated in the COA vs. AOA meta-analyses were compared using the Deming regression method^35^. The intercept was set to be 0; the slope estimated from the regression analysis is reported here.

**Supplementary Figure 12.**
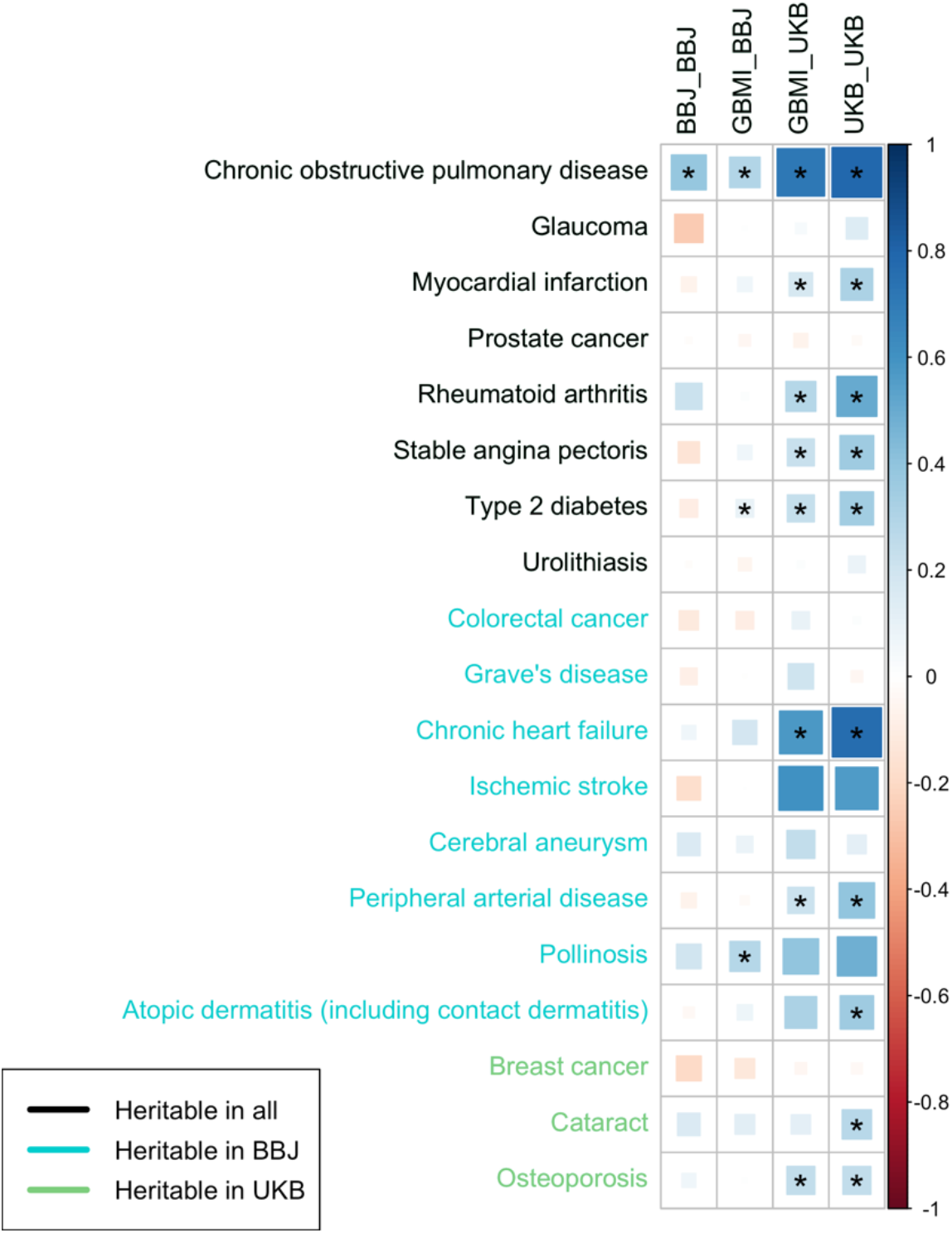
Genetic correlations between asthma and heritable diseases across UKBB and BBJ. Genetic correlations between asthma and disease endpoints that were significantly heritable in BBJ, UKBB EUR, or both. On x-axis: BBJ_BBJ = BBJ GWAS of asthma vs. BBJ GWAS of diseases on y-axis; GBMI_BBJ = GBMI-excluding-BBJ meta-analysis of asthma vs. BBJ GWAS of diseases on y-axis; GBMI_UKB = GBMI-excluding-UKB meta-analysis of asthma vs. UKB GWAS of diseases (EUR only) on y-axis; UKB_UKB = UKB GWAS of asthma vs. UKB GWAS of diseases (EUR only) on y-axis

### Supplementary Table Legends

**STable 1:** Description of 22 biobanks in GBMI that contributed summary statistics for asthma meta-analysis with sample size, age, ancestry, and recruitment strategy information.

**STable 2:** 179 index variants of top loci discovered by GBMI asthma meta-analysis with annotations for nearby genes, missense variants, and fine-mapped cis-eQTLs.

**STable 3:** Description of asthma phenotype definition used by each biobank.

**STable 4:** 122 of the 179 index variants present or with tagging variant in the TAGC study with effect sizes and p-values from GBMI and TAGC meta-analyses.

**STable 5:** Correlations of SNP effects of the 179 index variants between each biobank and the corresponding leave-that-biobank-out meta-analysis.

**STable 6:** Additional genome-wide significant loci identified by MR-MEGA not identified by fixed-effects inverse-variance weighted meta-analysis.

**STable 7:** Heterogeneity p-values, computed from Cochran’s Q statistic, for index variants using effect sizes from the AFR, AMR, EAS, EUR, and CSA meta-analyses.

**STable 8:** Loci not significant in the biobank meta-analysis with only EUR ancestry participants in GBMI.

**STable 9:** PRS accuracy results (R2 on liability scale) across target cohorts, including the downsampled EUR target cohort with 1k individuals.

**STable 10:** Description of discovery data used in PRS-CSx and PRS-CS analyses.

**STable 11:** Genome-wide significant loci from COA and AOA meta-analyses.

**STable 12:** Genetic correlations estimated by LDSC between GBMI leave-UKBB-out meta-analysis and UKBB EUR GWAS of significantly heritable phenotypes.

**STable 13:** Genetic correlations estimated by LDSC between GBMI asthma meta-analyses and UKBB and BBJ GWAS of several diseases.

**STable 14:** Genes prioritized by MAGMA using GBMI EAS, CSA, and EUR asthma meta-analyses. Genes with p-values < Bonferroni-corrected p-value thresholds are reported.

**STable 15:** Genes prioritized by MAGMA using GBMI EAS and EUR COPD meta-analyses. Genes with p-value < Bonferroni-corrected p-value thresholds are reported.

**STable 16:** Genes prioritized by DEPICT for asthma and COPD.

**STable 17:** Genes prioritized by PoPS for asthma and COPD. Genes with the highest 1% of PoPS scores are reported.

**STable 18:** Gene sets enriched by MAGMA for asthma. Gene sets with FDR < 0.05 are reported.

**STable 19:** Gene sets enriched by MAGMA for COPD. Gene sets with FDR < 0.05 are reported.

## Notes

### Author Declarations

Please refer to Supplementary Note for information regarding individual studies involved in the Global Biobank Meta-analysis Initiative.

### Summary of Updates

We added a new section "Childhood-onset (COA) and adult-onset (AOA) asthma are highly genetically correlated" and re-organized a previous section into two sections: "Asthma and COPD have shared and distinct biological processes" and "Genetic overlap between asthma and other diseases." Supplemental files are updated.

## References

1. Thomsen, S. F., van der Sluis, S., Kyvik, K. O., Skytthe, A. & Backer, V. Estimates of asthma heritability in a large twin sample. Clin. Exp. Allergy 40, 1054–1061 (2010).

2. Hernandez-Pacheco, N., Pino-Yanes, M. & Flores, C. Genomic Predictors of Asthma Phenotypes and Treatment Response. Front Pediatr 7, 6 (2019).

3. Moffatt, M. F. et al. Genetic variants regulating ORMDL3 expression contribute to the risk of childhood asthma. Nature 448, 470–473 (2007).

4. Moffatt, M. F. et al. A large-scale, consortium-based genomewide association study of asthma. N. Engl. J. Med. 363, 1211–1221 (2010).

5. Ferreira, M. A. R. et al. Identification of IL6R and chromosome 11q13.5 as risk loci for asthma. Lancet 378, 1006–1014 (2011).

6. El-Husseini, Z. W., Gosens, R., Dekker, F. & Koppelman, G. H. The genetics of asthma and the promise of genomics-guided drug target discovery. Lancet Respir Med 8, 1045–1056 (2020).

7. Han, Y. et al. Genome-wide analysis highlights contribution of immune system pathways to the genetic architecture of asthma. Nat. Commun. 11, 1776 (2020).

8. Ober, C. et al. Meta-analysis of genome-wide association studies of asthma in ethnically diverse North American populations. Nature Genetics vol. 43 887–892 (2011).

9. Demenais, F. et al. Multiancestry association study identifies new asthma risk loci that colocalize with immune-cell enhancer marks. Nat. Genet. 50, 42–53 (2018).

10. Sembajwe, G. et al. National income, self-reported wheezing and asthma diagnosis from the World Health Survey. Eur. Respir. J. 35, 279–286 (2010).

11. Asher, M. I., García-Marcos, L., Pearce, N. E. & Strachan, D. P. Trends in worldwide asthma prevalence. Eur. Respir. J. 56, (2020).

12. Akinbami, L. J. et al. Trends in asthma prevalence, health care use, and mortality in the United States, 2001-2010. NCHS Data Brief 1–8 (2012).

13. Dharmage, S. C., Perret, J. L. & Custovic, A. Epidemiology of Asthma in Children and Adults. Front Pediatr 7, 246 (2019).

14. Borish, L. & Culp, J. A. Asthma: a syndrome composed of heterogeneous diseases. Ann. Allergy Asthma Immunol. 101, 1–8; quiz 8–11, 50 (2008).

15. Kuruvilla, M. E., Lee, F. E.-H. & Lee, G. B. Understanding Asthma Phenotypes, Endotypes, and Mechanisms of Disease. Clin. Rev. Allergy Immunol. 56, 219–233 (2019).

16. Maselli, D. J. & Hanania, N. A. Asthma COPD overlap: Impact of associated comorbidities. Pulm. Pharmacol. Ther. 52, 27–31 (2018).

17. Postma, D. S. & Rabe, K. F. The Asthma–COPD Overlap Syndrome. N. Engl. J. Med. 373, 1241–1249 (2015).

18. Ferreira, M. A. R. et al. Genome-wide association analysis identifies 11 risk variants associated with the asthma with hay fever phenotype. J. Allergy Clin. Immunol. 133, 1564–1571 (2014).

19. Zhu, Z., Hasegawa, K., Camargo, C. A. & Liang, L. Investigating asthma heterogeneity through shared and distinct genetics: Insights from genome-wide cross-trait analysis. J. Allergy Clin. Immunol. 147, 796–807 (2021).

20. Zhu, Z. et al. Shared genetic and experimental links between obesity-related traits and asthma subtypes in UK Biobank. J. Allergy Clin. Immunol. 145, 537–549 (2020).

21. Zhu, Z. et al. A genome-wide cross-trait analysis from UK Biobank highlights the shared genetic architecture of asthma and allergic diseases. Nat. Genet. 50, 857–864 (2018).

22. Zhu, Z. et al. Shared genetics of asthma and mental health disorders: a large-scale genome-wide cross-trait analysis. Eur. Respir. J. 54, (2019).

23. Van Wonderen, K. E. et al. Different definitions in childhood asthma: how dependable is the dependent variable? Eur. Respir. J. 36, 48–56 (2010).

24. Colicino, S., Munblit, D., Minelli, C., Custovic, A. & Cullinan, P. Validation of childhood asthma predictive tools: A systematic review. Clin. Exp. Allergy 49, 410–418 (2019).

25. Lambert, S. A., Abraham, G. & Inouye, M. Towards clinical utility of polygenic risk scores. Hum. Mol. Genet. 28, R133–R142 (2019).

26. Lewis, C. M. & Vassos, E. Polygenic risk scores: from research tools to clinical instruments. Genome Med. 12, 44 (2020).

27. Chatterjee, N., Shi, J. & García-Closas, M. Developing and evaluating polygenic risk prediction models for stratified disease prevention. Nat. Rev. Genet. 17, 392–406 (2016).

28. Dijk, F. N. et al. Genetic risk scores do not improve asthma prediction in childhood. J. Allergy Clin. Immunol. 144, 857–860.e7 (2019).

29. Kothalawala, D. M. et al. Integration of Genomic Risk Scores to Improve the Prediction of Childhood Asthma Diagnosis. J Pers Med 12, (2022).

30. Belsky, D. W. et al. Polygenic risk and the development and course of asthma: an analysis of data from a four-decade longitudinal study. Lancet Respir Med 1, 453–461 (2013).

31. Sordillo, J. E. et al. A polygenic risk score for asthma in a large racially diverse population. Clin. Exp. Allergy 51, 1410–1420 (2021).

32. Namjou, B. et al. Multiancestral polygenic risk score for pediatric asthma. J. Allergy Clin. Immunol. (2022) doi:10.1016/j.jaci.2022.03.035.

33. Zhou, W. et al. Global Biobank Meta-analysis Initiative: powering genetic discovery across human diseases. medRxiv (2021) doi:10.1101/2021.11.19.21266436.

34. 1000 Genomes Project Consortium et al. A global reference for human genetic variation. Nature 526, 68–74 (2015).

35. Deming, W. E. Statistical adjustment of data. 261, (1943).

36. Jefferson, J. A. & Shankland, S. J. Familial nephrotic syndrome: PLCE1 enters the fray. Nephrol. Dial. Transplant 22, 1849–1852 (2007).

37. Riar, S. S. et al. Prevalence of Asthma and Allergies and Risk of Relapse in Childhood Nephrotic Syndrome: Insight into Nephrotic Syndrome Cohort. J. Pediatr. 208, 251–257.e1 (2019).

38. UK Biobank — Neale lab. http://www.nealelab.is/uk-biobank/.

39. Loo, T. H. et al. The mammalian LINC complex component SUN1 regulates muscle regeneration by modulating drosha activity. Elife 8, (2019).

40. Chen, M.-H. et al. Trans-ethnic and Ancestry-Specific Blood-Cell Genetics in 746,667 Individuals from 5 Global Populations. Cell 182, 1198–1213.e14 (2020).

41. Dalakas, M. C. & Spaeth, P. J. The importance of FcRn in neuro-immunotherapies: From IgG catabolism, FCGRT gene polymorphisms, IVIg dosing and efficiency to specific FcRn inhibitors. Ther. Adv. Neurol. Disord. 14, 1756286421997381 (2021).

42. Nebert, D. W. & Liu, Z. SLC39A8 gene encoding a metal ion transporter: discovery and bench to bedside. Hum. Genomics 13, 51 (2019).

43. Schizophrenia Working Group of the Psychiatric Genomics Consortium. Biological insights from 108 schizophrenia-associated genetic loci. Nature 511, 421–427 (2014).

44. Li, D. et al. A Pleiotropic Missense Variant in SLC39A8 Is Associated With Crohn’s Disease and Human Gut Microbiome Composition. Gastroenterology 151, 724–732 (2016).

45. Huang, H. et al. Fine-mapping inflammatory bowel disease loci to single-variant resolution. Nature 547, 173–178 (2017).

46. Speliotes, E. K. et al. Association analyses of 249,796 individuals reveal 18 new loci associated with body mass index. Nat. Genet. 42, 937–948 (2010).

47. Pickrell, J. K. et al. Detection and interpretation of shared genetic influences on 42 human traits. Nat. Genet. 48, 709–717 (2016).

48. International Consortium for Blood Pressure Genome-Wide Association Studies et al. Genetic variants in novel pathways influence blood pressure and cardiovascular disease risk. Nature 478, 103–109 (2011).

49. Nakata, T. et al. A missense variant in SLC39A8 confers risk for Crohn’s disease by disrupting manganese homeostasis and intestinal barrier integrity. Proc. Natl. Acad. Sci. U. S. A. 117, 28930–28938 (2020).

50. Qi, T. et al. Identifying gene targets for brain-related traits using transcriptomic and methylomic data from blood. Nat. Commun. 9, 2282 (2018).

51. Mägi, R. et al. Trans-ethnic meta-regression of genome-wide association studies accounting for ancestry increases power for discovery and improves fine-mapping resolution. Hum. Mol. Genet. 26, 3639–3650 (2017).

52. Massoud, A. H. et al. An asthma-associated IL4R variant exacerbates airway inflammation by promoting conversion of regulatory T cells to TH17-like cells. Nat. Med. 22, 1013–1022 (2016).

53. Kousha, A. et al. Interleukin 4 gene polymorphism (−589C/T) and the risk of asthma: a meta-analysis and met-regression based on 55 studies. BMC Immunol. 21, 55 (2020).

54. Nie, W., Zhu, Z., Pan, X. & Xiu, Q. The interleukin-4 -589C/T polymorphism and the risk of asthma: A meta-analysis including 7345 cases and 7819 controls. Gene vol. 520 22–29 (2013).

55. Battle, N. C. et al. Ethnicity-specific Gene–Gene Interaction between IL-13 and IL-4Rα among African Americans with Asthma. Am. J. Respir. Crit. Care Med. 175, 881–887 (2007).

56. Wang, Y. et al. Global biobank analyses provide lessons for computing polygenic risk scores across diverse cohorts. medRxiv (2021).

57. Ge, T., Chen, C.-Y., Ni, Y., Feng, Y.-C. A. & Smoller, J. W. Polygenic prediction via Bayesian regression and continuous shrinkage priors. Nat. Commun. 10, 1776 (2019).

58. Ruan, Y. et al. Improving Polygenic Prediction in Ancestrally Diverse Populations. bioRxiv (2021) doi:10.1101/2020.12.27.20248738.

59. Ferreira, M. A. R. et al. Genetic Architectures of Childhood-and Adult-Onset Asthma Are Partly Distinct. Am. J. Hum. Genet. 104, 665–684 (2019).

60. Pividori, M., Schoettler, N., Nicolae, D. L., Ober, C. & Im, H. K. Shared and distinct genetic risk factors for childhood-onset and adult-onset asthma: genome-wide and transcriptome-wide studies. Lancet Respir Med 7, 509–522 (2019).

61. Marco, R. de et al. The Coexistence of Asthma and Chronic Obstructive Pulmonary Disease (COPD): Prevalence and Risk Factors in Young, Middle-aged and Elderly People from the General Population. PLoS ONE vol. 8 e62985 (2013).

62. Madore, A.-M. et al. Analysis of GWAS-nominated loci for lung cancer and COPD revealed a new asthma locus. BMC Pulm. Med. 22, 155 (2022).

63. Hobbs, B. D. et al. Genetic loci associated with chronic obstructive pulmonary disease overlap with loci for lung function and pulmonary fibrosis. Nat. Genet. 49, 426–432 (2017).

64. Sakornsakolpat, P. et al. Genetic landscape of chronic obstructive pulmonary disease identifies heterogeneous cell-type and phenotype associations. Nat. Genet. 51, 494–505 (2019).

65. de Leeuw, C. A., Mooij, J. M., Heskes, T. & Posthuma, D. MAGMA: generalized gene-set analysis of GWAS data. PLoS Comput. Biol. 11, e1004219 (2015).

66. Pers, T. H. et al. Biological interpretation of genome-wide association studies using predicted gene functions. Nat. Commun. 6, 5890 (2015).

67. Weeks, E. M. et al. Leveraging polygenic enrichments of gene features to predict genes underlying complex traits and diseases. bioRxiv (2020) doi:10.1101/2020.09.08.20190561.

68. Subramanian, A. et al. Gene set enrichment analysis: a knowledge-based approach for interpreting genome-wide expression profiles. Proc. Natl. Acad. Sci. U. S. A. 102, 15545–15550 (2005).

69. Leynaert, B., Neukirch, F., Demoly, P. & Bousquet, J. Epidemiologic evidence for asthma and rhinitis comorbidity. J. Allergy Clin. Immunol. 106, S201–5 (2000).

70. Van Lieshout, R. J. & Macqueen, G. Psychological factors in asthma. Allergy Asthma Clin. Immunol. 4, 12–28 (2008).

71. Miethe, S., Karsonova, A., Karaulov, A. & Renz, H. Obesity and asthma. J. Allergy Clin. Immunol. 146, 685–693 (2020).

72. Guo, H., An, J. & Yu, Z. Identifying Shared Risk Genes for Asthma, Hay Fever, and Eczema by Multi-Trait and Multiomic Association Analyses. Front. Genet. 11, 270 (2020).

73. Kim, S. Y., Min, C., Oh, D. J. & Choi, H. G. Increased risk of asthma in patients with rheumatoid arthritis: A longitudinal follow-up study using a national sample cohort. Sci. Rep. 9, 6957 (2019).

74. Shen, T.-C., Lin, C.-L., Wei, C.-C., Tu, C.-Y. & Li, Y.-F. The risk of asthma in rheumatoid arthritis: a population-based cohort study. QJM 107, 435–442 (2014).

75. Luo, Y. et al. Rheumatoid arthritis is associated with increased in-hospital mortality in asthma exacerbations: a nationwide study. Clin. Rheumatol. 37, 1971–1976 (2018).

76. Jeong, H. E., Jung, S.-M. & Cho, S.-I. Association between Rheumatoid Arthritis and Respiratory Allergic Diseases in Korean Adults: A Propensity Score Matched Case-Control Study. Int. J. Rheumatol. 2018, 3798124 (2018).

77. Rolfes, M. C., Juhn, Y. J., Wi, C.-I. & Sheen, Y. H. Asthma and the Risk of Rheumatoid Arthritis: An Insight into the Heterogeneity and Phenotypes of Asthma. Tuberc. Respir. Dis. 80, 113–135 (2017).

78. Shirai, Y. et al. Multi-trait and cross-population genome-wide association studies across autoimmune and allergic diseases identify shared and distinct genetic component. Ann. Rheum. Dis. (2022) doi:10.1136/annrheumdis-2022-222460.

79. Ehrlich, S. F., Quesenberry, C. P., Jr, Van Den Eeden, S. K., Shan, J. & Ferrara, A. Patients diagnosed with diabetes are at increased risk for asthma, chronic obstructive pulmonary disease, pulmonary fibrosis, and pneumonia but not lung cancer. Diabetes Care 33, 55–60 (2010).

80. Thomsen, S. F., Duffy, D. L., Kyvik, K. O., Skytthe, A. & Backer, V. Risk of asthma in adult twins with type 2 diabetes and increased body mass index. Allergy 66, 562–568 (2011).

81. Torres, R. M., Souza, M. D. S., Coelho, A. C. C., de Mello, L. M. & Souza-Machado, C. Association between Asthma and Type 2 Diabetes Mellitus: Mechanisms and Impact on Asthma Control—A Literature Review. Can. Respir. J. 2021, (2021).

82. Sun, Y.-Q. et al. Adiposity and asthma in adults: a bidirectional Mendelian randomisation analysis of The HUNT Study. Thorax 75, 202–208 (2020).

83. Zhu, Z. et al. A large-scale genome-wide association analysis of lung function in the Chinese population identifies novel loci and highlights shared genetic aetiology with obesity. Eur. Respir. J. 58, (2021).

84. Tanaka, N. et al. Eight novel susceptibility loci and putative causal variants in atopic dermatitis. J. Allergy Clin. Immunol. 148, 1293–1306 (2021).

85. Park, J. et al. Predicting allergic diseases in children using genome-wide association study (GWAS) data and family history. World Allergy Organ. J. 14, 100539 (2021).

86. Majara, L. et al. Low generalizability of polygenic scores in African populations due to genetic and environmental diversity. Cold Spring Harbor Laboratory 2021.01.12.426453 (2021) doi:10.1101/2021.01.12.426453.

87. Kumbhare, S., Pleasants, R., Ohar, J. A. & Strange, C. Characteristics and Prevalence of Asthma/Chronic Obstructive Pulmonary Disease Overlap in the United States. Ann. Am. Thorac. Soc. 13, 803–810 (2016).

88. Hosseini, M., Almasi-Hashiani, A., Sepidarkish, M. & Maroufizadeh, S. Global prevalence of asthma-COPD overlap (ACO) in the general population: a systematic review and meta-analysis. Respir. Res. 20, 229 (2019).

89. Akmatov, M. K. et al. Comorbidity profile of patients with concurrent diagnoses of asthma and COPD in Germany. Sci. Rep. 10, 17945 (2020).

90. Munafò, M. R., Tilling, K., Taylor, A. E., Evans, D. M. & Davey Smith, G. Collider scope: when selection bias can substantially influence observed associations. Int. J. Epidemiol. 47, 226–235 (2018).

91. Griffith, G. J. et al. Collider bias undermines our understanding of COVID-19 disease risk and severity. Nat. Commun. 11, 5749 (2020).

92. Bulik-Sullivan, B. K. et al. LD Score regression distinguishes confounding from polygenicity in genome-wide association studies. Nat. Genet. 47, 291–295 (2015).

93. Denny, J. C. et al. Systematic comparison of phenome-wide association study of electronic medical record data and genome-wide association study data. Nat. Biotechnol. 31, 1102–1110 (2013).

94. Cann, H. M. et al. A human genome diversity cell line panel. Science 296, 261–262 (2002).

95. Karczewski, K. J. et al. Author Correction: The mutational constraint spectrum quantified from variation in 141,456 humans. Nature 590, E53 (2021).

96. Buniello, A. et al. The NHGRI-EBI GWAS Catalog of published genome-wide association studies, targeted arrays and summary statistics 2019. Nucleic Acids Res. 47, D1005–D1012 (2019).

97. Wang, K., Li, M. & Hakonarson, H. ANNOVAR: functional annotation of genetic variants from high-throughput sequencing data. Nucleic Acids Res. 38, e164 (2010).

98. McLaren, W. et al. The Ensembl Variant Effect Predictor. Genome Biol. 17, 122 (2016).

99. Aguet, F. et al. The GTEx Consortium atlas of genetic regulatory effects across human tissues. Science (2020).

100. Kerimov, N. et al. A compendium of uniformly processed human gene expression and splicing quantitative trait loci. Nat. Genet. 53, 1290–1299 (2021).

101. Wang, G., Sarkar, A., Carbonetto, P. & Stephens, M. A simple new approach to variable selection in regression, with application to genetic fine mapping. J. R. Stat. Soc. Series B Stat. Methodol. 82, 1273–1300 (2020).

102. Ulirsch, J. C. & Kanai, M. An annotated atlas of causal variants underlying complex traits and gene expression.

103. Kanai, M. et al. Insights from complex trait fine-mapping across diverse populations. bioRxiv (2021) doi:10.1101/2021.09.03.21262975.

104. Higgins, J. P. T. & Thompson, S. G. Quantifying heterogeneity in a meta-analysis. Stat. Med. 21, 1539–1558 (2002).

105. Boughton, A. P. et al. LocusZoom.js: Interactive and embeddable visualization of genetic association study results. Bioinformatics (2021) doi:10.1093/bioinformatics/btab186.

106. Purcell, S. & Chang, C. PLINK 1.9. https://www.cog-genomics.org/plink/1.9/.

107. Chang, C. C. et al. Second-generation PLINK: rising to the challenge of larger and richer datasets. Gigascience 4, 7 (2015).

108. Zhou, W. et al. Efficiently controlling for case-control imbalance and sample relatedness in large-scale genetic association studies. Nat. Genet. 50, 1335–1341 (2018).

109. Finucane, H. K. et al. Partitioning heritability by functional annotation using genome-wide association summary statistics. Nat. Genet. 47, 1228–1235 (2015).

110. Finucane, H. K. et al. Heritability enrichment of specifically expressed genes identifies disease-relevant tissues and cell types. Nat. Genet. 50, 621–629 (2018).

111. Pan UKBB. https://pan.ukbb.broadinstitute.org/ https://pan.ukbb.broadinstitute.org/.

112. Akiyama, M. et al. Genome-wide association study identifies 112 new loci for body mass index in the Japanese population. Nat. Genet. 49, 1458–1467 (2017).

113. Kanai, M. et al. Genetic analysis of quantitative traits in the Japanese population links cell types to complex human diseases. Nat. Genet. 50, 390–400 (2018).

114. Ishigaki, K. et al. Large-scale genome-wide association study in a Japanese population identifies novel susceptibility loci across different diseases. Nat. Genet. 52, 669–679 (2020).

115. Wei, T. & Simko, V. R package ‘corrplot’: Visualization of a Correlation Matrix. https://github.com/taiyun/corrplot (2021).

116. Therneau, T. deming: Deming, Theil-Sen, Passing-Bablock and Total Least Squares Regression. https://CRAN.R-project.org/package=deming (2018).

